# Comparative analysis of the outcomes of COVID-19 between patients infected with SARS-CoV-2 Omicron and Delta variants: a retrospective cohort study

**DOI:** 10.1101/2022.04.30.22274532

**Authors:** Gunadi, Mohamad Saifudin Hakim, Hendra Wibawa, Khanza Adzkia Vujira, Dyah Ayu Puspitarani, Endah Supriyati, Ika Trisnawati, Kristy Iskandar, Riat El Khair, Afiahayati, Siswanto, Yunika Puspadewi, Irene, Sri Handayani Irianingsih, Edwin Widyanto Daniwijaya, Dwi Aris Agung Nugrahaningsih, Gita Christy Gabriela, Esensi Tarian Geometri, Laudria Stella Eryvinka, Fadila Dyah Trie Utami, Edita Mayda Devana, Lanang Aditama, Nathania Christi Putri Kinasih, Verrell Christopher Amadeus, Yekti Hediningsih, Nur Rahmi Ananda, Eggi Arguni, Titik Nuryastuti, Tri Wibawa, the Yogyakarta-Central Java COVID-19 study group

## Abstract

**Background:** The SARS-CoV-2 Omicron variant has replaced the previously dominant Delta variant because of high transmissibility. It is responsible for the current increase in the COVID-19 infectivity rate worldwide. However, studies on the impact of the Omicron variant on the severity of COVID-19 are still limited in developing countries. Here, we compared the outcomes of patients infected with SARS-CoV-2 Omicron and Delta variants and associated with prognostic factors, including age, sex, comorbidities, and smoking.

**Methods:** We involved 352 patients, 139 with the Omicron variant and 213 with the Delta variant. The whole-genome sequences of SARS-CoV-2 were conducted using the Illumina MiSeq next-generation sequencer.

**Results:** Ct value and mean age of COVID-19 patients were not significantly different between both groups (Delta: 20.35 ± 4.07 *vs*. Omicron: 20.62 ± 3.75; *p*=0.540; and Delta: 36.52 ± 21.24 *vs*. Omicron: 39.10 ± 21.24; *p*=0.266, respectively). Patients infected with Omicron and Delta variants showed similar hospitalization (*p*=0.433) and mortality rates (*p*=0.565). Multivariate analysis showed that older age (≥65 years) had higher risk for hospitalization (OR=3.67 [95% CI=1.22-10.94]; *p*=0.019) and fatalities (OR=3.93 [95% CI=1.35-11.42]; *p*=0.012). In addition, patients with cardiovascular disease had higher risk for hospitalization (OR=5.27 [95% CI=1.07-25.97]; *p*=0.041), whereas patients with diabetes revealed higher risk for fatalities (OR=9.39 [95% CI=3.30-26.72]; *p*=<0.001).

**Conclusions:** Our study shows that patients infected with Omicron and Delta variants reveal similar clinical outcomes, including hospitalization and mortality. In addition, our findings further confirm that older age, cardiovascular disease, and diabetes are strong prognostic factors for the outcomes of COVID-19 patients.

## Introduction

Severe acute respiratory syndrome coronavirus 2 (SARS-CoV-2) is the causative agent of the ongoing global pandemic of Coronavirus disease 2019 (COVID-19). As RNA viruses, SARS-CoV-2 is undergoing continuous mutation and evolution, giving rise to novel variants with the selective fitness advantage [1]. These variants have different characteristics due to their unique sets of mutations, including transmission rate, immune escape, and clinical severity. Variants with sufficient evidence of increased transmissibility, more severe diseases, or reduced vaccine or antiviral drug effectiveness have been designated a variant of concern (VOC). Therefore, these variants continuously impact both global health and the economy, with millions of people infected, hospitalized, and dead [2,3].

The Omicron variant (B.1.1.529) was first identified in a sample collected in Botswana on November 11, 2021. However, it was first reported by South Africa on November 24, 2021. Only within two days, the Omicron was classified as a variant of concern (VOC) by the World Health Organization on November 26, 2021 [4,5]. In Indonesia, the surge of COVID-19 cases due to the Omicron variant occurred from late January until February 2022. Subsequently, the Omicron variant was found as the most frequently detected VOC compared to the previously dominating Delta variant [6]. SARS-CoV-2 genomic surveillance in Indonesia is continuously conducted to monitor circulating SARS-CoV-2 variants.

Because of high transmissibility, the SARS-CoV-2 Omicron variant has replaced the previous dominant variant, i.e., the Delta variant. It was responsible for the current increase in the COVID-19 infectivity rate worldwide, including in Indonesia [7-9]. However, studies on the impact of the Omicron variant on the clinical severity of COVID-19 are still limited in developed countries [10-12]. Several studies in developed countries showed a lower clinical severity of Omicron-infected patients than Delta-infected patients [10-12]. Several factors have been associated with the clinical severity of infection, including vaccination status and the previous SARS-CoV-2 infection. In addition, we and others have shown that several prognostic factors have been associated with the outcomes of COVID-19 [13-17]. Therefore, to further comprehend the impact of the Omicron wave in developing countries, we performed a comparative analysis of the outcomes of COVID-19 patients infected with Delta and Omicron variants and associated them with several prognostic factors, including age, sex, comorbidities, and smoking. Our study thus contributes to informing the public health response against the emerging SARS-CoV-2 variants.

## Material and Methods

### Subjects

We used data from 352 patients with COVID-19 from Yogyakarta and Central Java provinces, Indonesia, from May 2021 to February 2022, consisting of 164 males and 188 females. The diagnosis of COVID-19 was defined according to PCR results for SARS-CoV-2. We included all patients with the PCR’s Ct value of less than 30 for further whole-genome sequencing. Outcomes of patients with COVID-19 were hospitalization and mortality. The Medical and Health Research Ethics Committee of the Faculty of Medicine, Public Health and Nursing, Universitas Gadjah Mada, approved our study (KE/FK/0563/EC/2020).

### Whole-genome sequencing of SARS-CoV-2

We collected all samples from nasopharyngeal swabs of outpatient or hospitalized patients with COVID-19 from May 2021 to February 2022. Subsequently, samples were sent to COVID-19 Testing Laboratory Network in Yogyakarta for PCR and to Balai Besar Veteriner Wates and Balai Besar Teknik Kesehatan Lingkungan dan Pengendalian Penyakit Yogyakarta for whole-genome sequencing using MiSeq Illumina Platform.

We conducted whole-genome sequencing (WGS) of SARS-CoV-2 for all samples with PCR’s Ct value of less than 30. As described in our previous studies [13,14,18,119], single-stranded cDNA was synthesized from total RNA extracted from the samples of COVID-19 patients by SuperScript™ III First-Strand Synthesis System (Thermo Fisher Scientific, MA, United States). Subsequently, the second strand was synthesized by COVID-19 ARTIC v3 primer pool design by SARS-CoV-2 ARTIC Network using Phusion™ High-Fidelity DNA Polymerase (Thermo Fisher Scientific, MA, United States). The library preparations were performed using the Illumina DNA Prep (Illumina, California, United States). The Illumina MiSeq next-generation sequencer was used to perform the whole genome sequences of SARS-CoV-2. The genomes of our samples were assembled and mapped into the reference genome from Wuhan, China (hCoV-19/Wuhan/Hu-1/2019, GenBank accession number: NC_045512.2) using Burrow-Wheeler Aligner (BWA) algorithm embedded in UGENE v. 1.30 [20].

### Phylogenetic study

A dataset of 392 available SARS-CoV-2 genomes was extracted from GISAID from our region and others (Acknowledgment Table is provided in Supp. Table 1) to reconstruct the phylogenetic tree. This included SARS-CoV-2 genomes from 352 virus samples collected from our study in Central Java and Yogyakarta Special Regions provinces during Delta and Omicron variant infection waves in 2021 and 2022 and 40 virus genomes of other SARS-CoV-2 variants that previously circulated in Indonesia (B.1.1.7/Alpha, B.1.466.2, B.36, and B.1.470). Firstly, a multiple nucleotide sequence alignment was performed using the MAFFT program version 7 (https://mafft.cbrc.jp/alignment/server/). The neighbor-joining statistical method with 1,000 bootstrap replications [21,22] was used to construct a phylogenetic tree from 29.409 nucleotide length of the open reading frame (ORF) of SARS-CoV-2, followed by computation of the evolutionary distances and model of the rate variation among sites by the Kimura 2-parameter method and the gamma distribution with estimated shape parameter (α) for the dataset, respectively [23]. The DAMBE version 7 [24] was utilized to calculate the estimation of the α gamma distribution, MEGA version 10 (MEGA X) [25] for phylogenetic reconstruction, and followed by tree visualization in FigTree (http://tree.bio.ed.ac.uk/software/FigTree/) to using a Newick tree output from MEGA X.

### Prognostic variables

We associated the outcomes of COVID-19 patients with the following prognostic variables: sex; age; comorbidities, including obesity, diabetes, hypertension, cardiovascular disease, and chronic kidney disease; and smoking. According to the previous reports, those prognostic factors were selected [13-17].

### Statistical analysis

The data were presented as mean ± SD and frequency (percentage). We determined the normality of the continuous variables by the Kolmogorov-Smirnov test. We excluded the missing or incomplete data from the final analysis. We used Chi-square or Fisher exact tests with a 95% confidence interval (CI) to find any significant association between variables and the outcomes of COVID-19 patients. Subsequently, we performed a multivariate analysis using a logistic regression test. We considered the *p*-value of <0.05 as significant. We conducted all statistical analyses by the IBM Statistical Package for the Social Sciences (SPSS) version 23 (Chicago, USA).

## Results

### Phylogenetic study

Phylogenetic analysis showed that about 60.5% (213 samples) of SARS-CoV-2 collected from Central Java and Yogyakarta provinces between May 2021 and February 2022 belonged to B.1.617.2-like (Delta variant), while 39.5% (139 samples) clustered in BA-like (Omicron variant) (Fig.1). The majority of Delta variants were AY.23 lineage (91.5%), whereas a small proportion was AY.24 lineage (8.5%). For the Omicron variant, BA.1 lineage was predominantly detected (77.7%), followed by BA.2 lineage (21.6%), and only one virus (0.7%) belonged to BA.3 lineage (hcov-19/Indonesia/YO-GS-22.02175/2022: EPI_ISL_9702414). In particular, to BA.1-like virus, SARS-CoV-2 viruses from Central Java and Yogyakarta that belonged to this lineage breached into three distinct groups consisting of BA.1.13, BA.1.14, BA.1.15 (Group I), BA.1.1, BA.1.17, BA.1.18 (Group II), and a unique cluster of BA.1.13 (Group III) (Figure 1).

**Figure 1.**
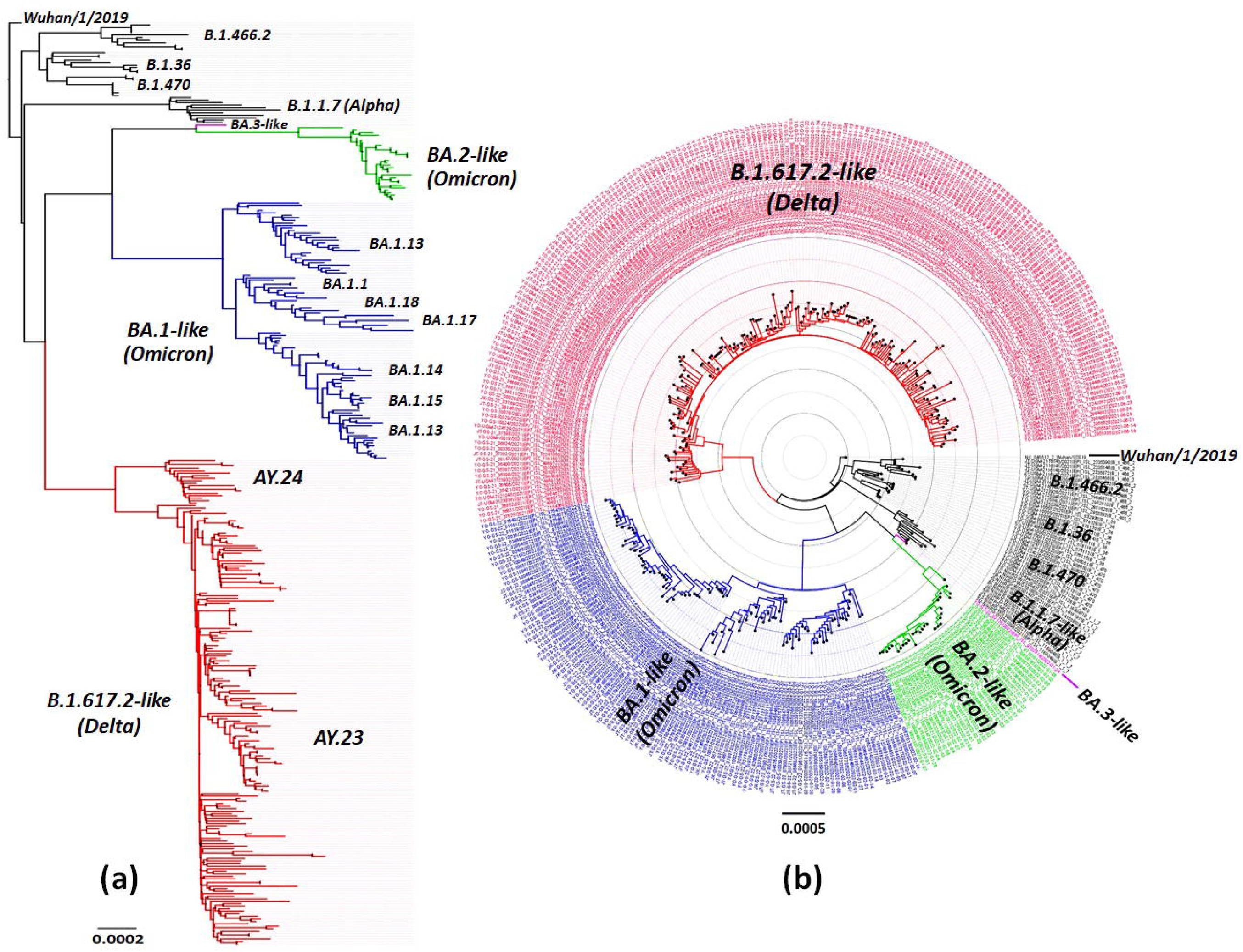
Phylogenetic analysis of Omicron and Delta variants of SARS-CoV-2 virus collected from Central Java and Yogyakarta regions from 2021-2022. Phylogenetic trees are displayed in rectangular (a) and polar (b) layouts. The evolutionary history was inferred using the Neighbor-Joining method [1] and computed using the Kimura 2-parameter [2] method with 1000 bootstrap replication in MEGA X [3,4]. The number of base substitutions per site (0.0001) is shown on the left of the rectangular tree, where the rate variation among sites was modeled with a gamma distribution. This analysis involved 392 nucleotide sequences, a total of 29.409 positions in the final dataset. Moreover, all ambiguous positions were removed for each sequence pair (pairwise deletion option). The Delta variant taxa are indicated in red, followed by Omicron-BA.1-like in blue, Omicron-BA.2-like in green, Omicron-BA.3-like in purple, and other variants previously circulating in Indonesia that are involved in this analysis are indicated in black.

### Clinical characteristics of our patients

Ct value and mean age were not significantly different between both groups (Delta: 20.35 ± 4.07 *vs*. Omicron: 20.62 ± 3.75; *p*=0.540; and Delta: 36.52 ± 21.24 *vs*. Omicron: 39.10 ± 21.24; *p*=0.266, respectively) (Table 1). All clinical characteristics of both groups were similar, except for the comorbidities of diabetes (*p*=0.022) and chronic kidney disease (*p*=0.019) (Table 1).

**Table 1.** Clinical characteristics of COVID-19 patients involved in this study.

| Characteristics | Total (N=352)<br>N (%); mean ± SD | Delta (N=213)<br>N (%); mean ± SD | Omicron (N=139)<br>N (%); mean ± SD | p-value |
| --- | --- | --- | --- | --- |
| Ct value | 20.46 ± 3.95 | 20.35 ± 4.07 | 20.62 ± 3.75 | 0.540 |
| Age (years) | 37.54 ± 21.25 | 36.52 ± 21.24 | 39.10 ± 21.24 | 0.266 |
| ≥65 | 37 (10.5) | 53 (24.9) | 28 (20.3) |  |
| 18 - <65 | 234 (66.5) | 140 (65.7) | 94 (68.1) |  |
| <18 | 81 (23) | 20 (9.4) | 17 (11.6) |  |
| Sex |  |  |  | 0.072 |
| Male | 164 (46.6) | 91 (42.7) | 73 (52.5) |  |
| Female | 188 (53.4) | 122 (57.2) | 66 (47.5) |  |
| Comorbidities |  |  |  |  |
| Obesity | 4 (1.1) | 4 (1.9) | 0 | 0.149 |
| Diabetes | 29 (8.2) | 23 (10.8) | 6 (4.3) | 0.022* |
| Hypertension | 36 (10.7) | 25 (11.7) | 12 (8.6) | 0.425 |
| Cardiovascular disease | 23 (6.9) | 12 (5.6) | 11 (7.9) | 0.464 |
| Chronic kidney disease | 7 (2.1) | 1 (0.5) | 6 (4.3) | 0.019* |
| Smoking | 17 (5.1) | 12 (5.6) | 5 (3.6) | 0.335 |

\*, $p < 0.05$ ***Prognostic factors for outcomes of patients with COVID-19***

### Prognostic factors for outcomes of patients with COVID-19

The hospitalization and mortality rates were not significantly different between patients infected with Omicron and Delta variants (Table 2). The older patients were less hospitalized than the younger subjects (*p*=<0.000), but they had a higher risk for mortality than younger ones (OR=6.91 [95%CI=2.84-16.86]; *p*=<0.000). Patients with diabetes, hypertension, cardiovascular disease, and chronic kidney disease showed a higher hospitalization risk. In addition, subjects with obesity, diabetes, hypertension, cardiovascular disease, and smoking revealed a higher risk for fatalities (Table 2).

**Table 2.** Prognostic factors for outcomes of COVID-19 patients

| Variables | Hospitalized<br>(N, %) | *p*-value | OR (95% CI) | Mortality (N,<br>%) | *p*-value | OR (95% CI) |
| --- | --- | --- | --- | --- | --- | --- |
| SARS-CoV-2 variant |  |  |  |  |  |  |
| ☐ Delta (N=213) | 111 (52.1) | 0.396 | 0.83 (0.54-1.27) | 19 (8.9) | 0.566 | 0.79 (0.36-1.76) |
| ☐ Omicron (N=139) | 66 (47.8) |  |  | 10 (7.2) |  |  |
| Age |  |  |  |  |  |  |
| ☐ <65-18 (N=234) | 124 (53.2) | Ref |  | 14 (6) | Ref |  |
| ☐ ≥65 (N=36) | 5 (13.9) | <0.000* | 0.14 (0.05-0.38) | 11 (30.6) | <0.000* | 6.91 (2.84-16.86) |
| ☐ <18 (N=82) | 37 (54.3) | 0.221 | 0.73 (0.44-1.2) | 4 (4.9) | 0.791 | 0.81 (0.26-2.52) |
| Sex |  |  |  |  |  |  |
| ☐ Female (N=188) | 96 (51.3) | 0.754 | 0.94 (0.62-1.42) | 13 (6.9) | 0.332 | 0.69 (0.32-1.48) |
| ☐ Male (N=164) | 81 (49.4) |  |  | 16 (9.8) |  |  |
| Comorbidity |  |  |  |  |  |  |
| a. Obesity |  |  |  |  |  |  |
| Yes (N=4) | 4 (100) | 0.123 | 9.10 (0.49-170.38) | 2 (50) | 0.035* | 11.90 (1.61-87.76) |
| No (N=348) | 173 (49.9) |  |  | 27 (7.8) |  |  |
| b. Diabetes |  |  |  |  |  |  |
| Yes (N=29) | 29 (100) | <0.000* | 69.73 (4.22-1150.98) | 13 (44.8) | <0.000* | 15.59(6.42-37.88) |
| No (N=323) | 148 (46) |  |  | 16 (5) |  |  |
| c. Hypertension |  |  |  |  |  |  |
| Yes (N=37) | 29 (80) | <0.000* | 4.09 (1.81-9.92) | 11 (31.4) | <0.000* | 7.28 (3.10-17.11) |
| No (N=315) | 149 (47.2) |  |  | 18 (5.7) |  |  |
| d. Cardiovascular disease |  |  |  |  |  |  |
| Yes (N=23) | 21 (91.3) | <0.000* | 11.64 (2.69-50.47) | 7 (30.4) | 0.001* | 6.11 (2.27-16.40) |
| No (N=329) | 156 (47.6) |  |  | 22 (6.7) |  |  |
| e. Chronic kidney disease |  |  |  |  |  |  |
| Yes (N=7) | 7 (100) | 0.015* | 2.02 (1.81-2.25) | 1 (14.3) | 0.56 | 1.89 (0.22-16.23) |
| No (N=345) | 170 (49.4) |  |  | 28 (8.1) |  |  |
| Smoking |  |  |  |  |  |  |
| ☐ Yes (N=17) | 9 (52.9) | 0.823 | 1.12 (0.42-2.97) | 4 (23.5) | 0.042* | 3.82 (1.16-12.57) |
| ☐ No (N=335) | 168 (50.3) |  |  | 25 (7.5) |  |  |

\*, significant ( $p<0.05$ ); CI, confidence interval; OR, odds ratio

### Multivariate analysis of prognostic factors

Subsequently, we conducted a multivariate analysis to determine the independent prognostic factors for the hospitalization and mortality of patients with COVID-19. The analysis revealed that older age (>65 years) had higher risk to be hospitalized (OR=3.67 [95% CI=1.23-10.94]; *p*=0.019) and died (OR=3.93 [95% CI=1.35-11.42]; *p*=0.012). In addition, patients with cardiovascular disease had higher risk to be hospitalized (OR=5.27 [95% CI=1.07-25.97]; *p*=0.041), whereas patients with diabetes revealed higher risk to be died (OR=9.39 [95% CI=3.30-26.72]; *p*=<0.001) (Table 3).

**Table 3.**
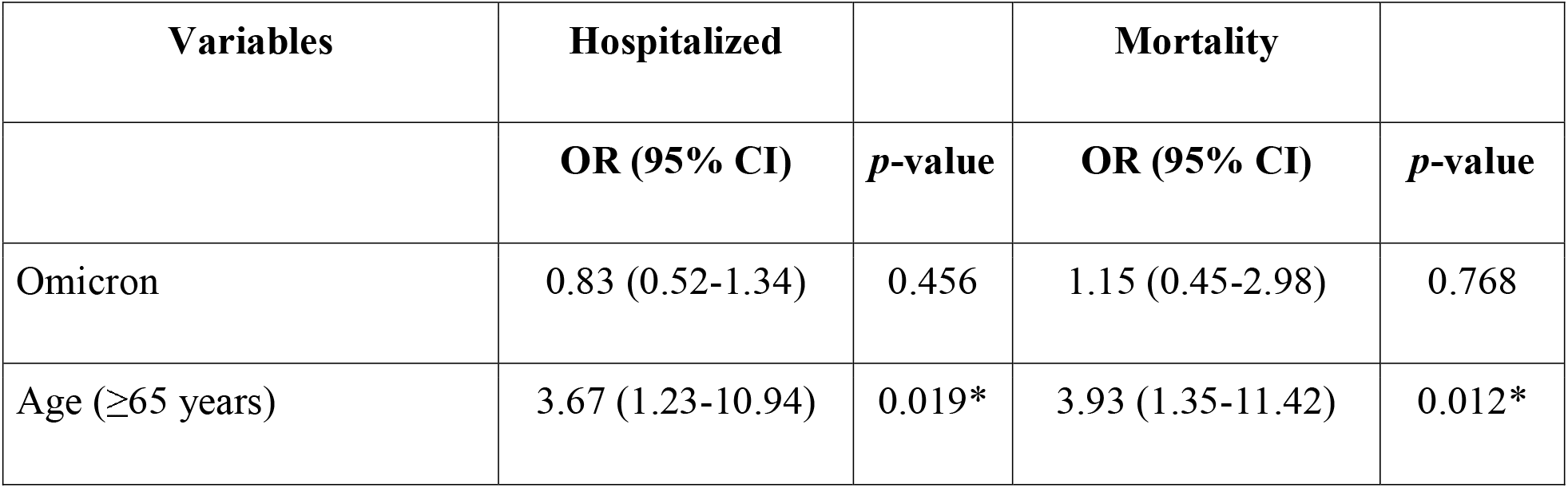
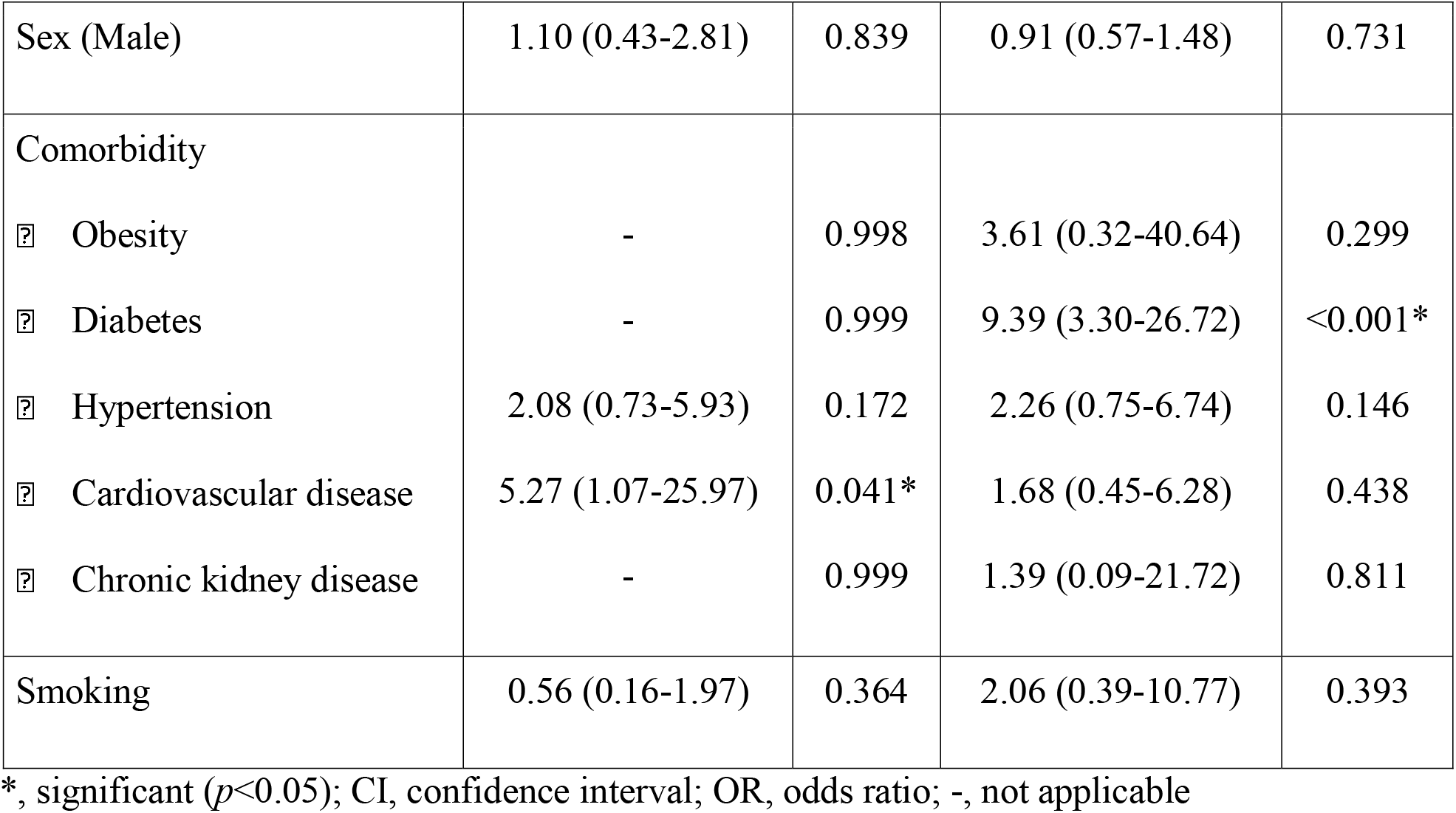
Multivariate analysis of prognostic factors for outcomes of patients with COVID-19.

| Variables | Hospitalized |  | Mortality |  |
| --- | --- | --- | --- | --- |
|  | OR (95% CI) | *p*-value | OR (95% CI) | *p*-value |
| Omicron | 0.83 (0.52-1.34) | 0.456 | 1.15 (0.45-2.98) | 0.768 |
| Age ($\geq 65$ years) | 3.67 (1.23-10.94) | 0.019* | 3.93 (1.35-11.42) | 0.012* |
| Sex (Male) | 1.10 (0.43-2.81) | 0.839 | 0.91 (0.57-1.48) | 0.731 |
| Comorbidity |  |  |  |  |
| ☐ Obesity | - | 0.998 | 3.61 (0.32-40.64) | 0.299 |
| ☐ Diabetes | - | 0.999 | 9.39 (3.30-26.72) | <0.001* |
| ☐ Hypertension | 2.08 (0.73-5.93) | 0.172 | 2.26 (0.75-6.74) | 0.146 |
| ☐ Cardiovascular disease | 5.27 (1.07-25.97) | 0.041* | 1.68 (0.45-6.28) | 0.438 |
| ☐ Chronic kidney disease | - | 0.999 | 1.39 (0.09-21.72) | 0.811 |
| Smoking | 0.56 (0.16-1.97) | 0.364 | 2.06 (0.39-10.77) | 0.393 |

\*, significant ( $p<0.05$ ); CI, confidence interval; OR, odds ratio; -, not applicable

## Discussion

Here, we show that the outcomes of patients infected with the Omicron variant are similar to patients infected with Delta variant regarding the hospitalization and mortality rates. Our findings were different from previous reports [10-12]. Nyberg *et al*. demonstrated that the outcomes of Omicron were significantly less severe than Delta and varied among ages [10]. Lewnard *et al*. showed that Omicron-infected patients had a lower risk of hospitalization, admission to the intensive care unit (ICU), use of ventilation, and death than Delta-infected patients. The differences were more significant in unvaccinated COVID-19 patients [11]. Importantly, Bouzid et al. indicated that Omicron patients had higher COVID-19 vaccination coverage [12]. Differences between our findings and previous reports might be affected by several variables, including host genetic background, public health measures, previous SARS-CoV-2 infections, and vaccination coverage [26-31,]. Unfortunately, incomplete data on vaccination status and previous SARS-CoV-2 infections of our patients hampered us to analyze further the impact of both variables on hospitalization and mortality in our patient cohort. Up to April 16, 2022, the vaccination coverage in Indonesia was 95.13% and 78.08% of the population for single and double doses, respectively [32]. In addition, the outcome of COVID-19 varies among the ethnic group [26-28]. Therefore, further study is essential to determine the association between host genetic risk alleles and the COVID-19 outcomes in our patients. Notably, while a previous study only analyzed patients admitted to the emergency department [12], we comprehensively analyzed the patients both in the hospital and community.

Previous studies showed that older age and comorbidities are significant prognostic factors for hospitalization and mortality [13-15,33,34]. Our study further provides evidence that older age, diabetes, and cardiovascular diseases as strong prognostic factors for the outcomes of COVID-19 patients. A recent systematic review showed that 50%, 20%, and 10% of older patients with COVID-19 had a severe illness, critical illness and died, respectively [35]. Several variables had attributed to the worse outcomes of COVID-19 in older patients, including associated comorbidities and delayed diagnosis due to atypical clinical manifestations [35]. Eighty percent of older patients with COVID-19 had a minimum of one comorbidity, such as hypertension, diabetes, and cardiovascular diseases [35]. Diabetes patients might have an uncontrolled immune response and increased ACE-2 receptors and furin during SARS-CoV-2 infection, while the use of antihypertension causes aberrant ACE-2 receptor expression in hypertension patients [36]. Both mechanisms lead to the severe illness of COVID-19 patients [36]. A recent report revealed that COVID-19 patients showed a higher risk of having cardiovascular diseases one month after acute infection [37]. They strongly suggest that the survival patients should be closely followed-up for the cardiovascular diseases after acute COVID-19 infection [37].

Children were more hospitalized during the Omicron surge than another variant surge [38]. The risk of hospitalization in children under four years during the Omicron surge was 5-times higher than during the Delta surge, particularly infants <6 months old [39]. However, the COVID-19 severity was not affected by age [39]. It is suggested to vaccinate any eligible subjects, including pregnant woman, family members, and their caregivers, to prevent children <4 years from getting COVID-19 infection [39]. Interestingly, maternal antibodies developed following vaccination can undergo transplacental transfer. Indeed, younger children can receive protection [39]. Our study did not show any difference in hospitalization and mortality rates between children and adults. Moreover, we grouped all pediatric populations from neonates, infants, young and older children into one group, i.e., <18 years.

A previous study showed that the Ct value of Omicron was significantly higher than Delta, implying that higher transmission of Omicron does not necessarily associate with its viral load [40]. Their findings might be affected by the vaccination and prior infection of COVID-19 due to the vaccination coverage and infection rate being higher during the Omicron than the Delta surge [40]. However, our study found that the Ct value was not significantly different between Omicron and Delta variants.

Although the clinical outcome of Omicron is less severe than Delta, it should be noted that Omicron has higher transmissibility and immune escape from previous COVID-19 infection and vaccination. These findings caused an extraordinary surge of COVID-19 globally and might affect the healthcare systems, including high absolute numbers of hospital admission and mortality rates [11]. Indeed, public health measures and vaccination are still crucial to control the COVID-19 spreading and decrease morbidity and mortality.

Several limitations are noted in our study including design of retrospective study, incomplete data on vaccination and previous infection of COVID-19, data were based on hospital admission, but no data from ICU admission unit and the use of mechanical ventilation, no follow-up data for patients after discharge from hospital, and incomparable sample size between Delta and Omicron variants.

## Conclusions

Our study shows that patients infected with Omicron and Delta variants reveal similar outcomes, including hospitalization and mortality. In addition, our findings further confirm that older age, cardiovascular disease, and diabetes are the strong prognostic factors for the outcomes of patients with COVID-19.

## Data Availability

All data produced in the present work are contained in the manuscript

https://www.gisaid.org/

## Data Availability Statement

All data generated or analyzed during this study are included in the submission. The sequence and metadata are shared through GISAID (www.gisaid.org).

## Ethics Statement

This study was approved by the Medical and Health Research Ethics Committee, Faculty of Medicine, Public Health and Nursing, Universitas Gadjah Mada/Dr. Sardjito Hospital, Yogyakarta, Indonesia (KE/FK/0563/EC/2020). The research has been performed following the Declaration of Helsinki. All participants or guardians signed written informed consent for participating in this study.

## Author Contributions

G and KI conceived the study. G drafted the manuscript, and MSH, HW, EA, and TW critically revised the manuscript for important intellectual content. KAV, DAP, GCG, ETG, LSE, FDTU, EMD, LA, NCPK, and VCA performed the library preparation and NGS. G, MSH, ES, IT, KI, REK, A, S, YP, I, SHI, EWD, DAAN, YH, NRA, and TN collected the data; and G, HW, KAV, and DAP analyzed the data. All authors have read and approved the manuscript and agreed to be accountable for all aspects of the work in ensuring that questions related to the accuracy or integrity of any part of the work are appropriately investigated and resolved.

## Acknowledgments

We thank the Collaborator Members of the Yogyakarta-Central Java COVID-19 study group: Eko Budiono, Heni Retnowulan, Sumardi, Bambang Sigit Riyanto, Munawar Gani, Satria Maulana, Ira Puspitawati, Osman Sianipar (Faculty of Medicine, Public Health and Nursing, Universitas Gadjah Mada/RSUP Dr. Sardjito), Lestari and Herdiyanto Mulyawan (Disease Investigation Center Wates/Balai Besar Besar Veteriner Wates, Yogyakarta), Indaryati and Havid Setyawan (Balai Besar Teknik Kesehatan Lingkungan dan Pengendalian Penyakit, Yogyakarta), Sri Fatmawati, Fatin Asfarina dan Sumartiningsih (Faculty of Medicine, Public Health and Nursing, Universitas Gadjah Mada), Safitriani and Muhammad Taufiq Soekarno (PT. Pandu Biosains). We gratefully acknowledge the authors, the originating and submitting laboratories for their sequence and metadata shared through GISAID. We also thank Besar Besar Veteriner Wates (Disease Investigation Center Wates) and Balai Besar Teknik Kesehatan Lingkungan dan Pengendalian Penyakit Yogyakarta, where the virus samples were sequenced using the NGS Illumina MiSeq instrument. All submitters of data may be contacted directly via www.gisaid.org. The Acknowledgments Table for GISAID is reported as Supplementary Table 1. The Indonesian Ministry of Health and Dr. Sardjito Hospital funded our study. The funders had no role in study design, data collection, and analysis, decision to publish, or manuscript preparation.

