## Supplementary Table 1 for "Comparative analysis of the outcomes of COVID-19 between patients infected with SARS-CoV-2 Omicron and Delta variants: a retrospective cohort study"

#### Accession ID Originating Laboratory Submitting Laboratory Authors

EPI_ISL_2943235	BBTKLPP Yogyakarta	Genetics Working Group (Pokja Genetik) Faculty of Medicine, Public Health and Nursing Universitas Gadjah Mada (FK-KMK UGM); Disease

Investigation Center Wates Ministry of Agriculture Indonesia; Department of Microbiology FK-KMK UGM; Laboratorium Diagnostik Yayasan Tahija World Mosquito Program (WMP) Yogyakarta Center for Tropical Medicine FK-KMK UGM; Integrated Research Center FK-KMK UGM; Department of Computer Science and Electronics FMIPA UGM; RSUP Dr. Sardjito

Afiahayati; Dwi AA Nugrahaningsih; Dwi Indaryati; Edwin W. Daniwijaya; Eggi Arguni; Endah Supriyati; Gunadi; Hana F Hanifin; Hendra Wibawa; Indarto Sulistiyono; Irene Tania; Kristy Iskandar; Ludhang P. Rizki; Marcellus; Mohamad S. Hakim; Nungki Anggorowati; Pramesti G Dewi; Siswanto; Titik Nuryastuti; Tri Wibawa

EPI_ISL_2964704, EPI_ISL_2964706

EPI_ISL_2964697, EPI_ISL_2964700

DINKES KAB. GROBOGAN	Genetics Working Group (Pokja Genetik) Faculty of Medicine, Public Health and Nursing Universitas Gadjah Mada (FK-KMK UGM); Disease Investigation Center Wates Ministry of Agriculture Indonesia; Department of Microbiology FK-KMK UGM; Laboratorium Diagnostik Yayasan Tahija World Mosquito Program (WMP) Yogyakarta Center for Tropical Medicine FK-KMK UGM; Integrated Research Center FK-KMK UGM; Department of

Computer Science and Electronics FMIPA UGM; RSUP Dr. Sardjito

DINKES KAB. JEPARA	Genetics Working Group (Pokja Genetik) Faculty of Medicine, Public Health and Nursing Universitas Gadjah Mada (FK-KMK UGM); Disease Investigation Center Wates Ministry of Agriculture Indonesia; Department of Microbiology FK-KMK UGM; Laboratorium Diagnostik Yayasan Tahija World Mosquito Program (WMP) Yogyakarta Center for Tropical Medicine FK-KMK UGM; Integrated Research Center FK-KMK UGM; Department of

Computer Science and Electronics FMIPA UGM; RSUP Dr. Sardjito

Afiahayati; Dwi AA Nugrahaningsih; Dwi Indaryati; Dyah A Puspitarani; Edwin W. Daniwijaya; Eggi Arguni; Endah Supriyati; Gunadi; Hana F Hanifin; Hendra Wibawa; Indarto Sulistiyono; Irene Tania; Kristy Iskandar; Ludhang P. Rizki; Marcellus; Mohamad S. Hakim; Nungki Anggorowati; Pramesti G Dewi; Siswanto; Susan Simanjaya; Titik Nuryastuti; Tri Wibawa

Afiahayati; Alvina A Setiawan; Cita S Amalia; Dwi AA Nugrahaningsih; Dwi Indaryati; Edwin W. Daniwijaya; Eggi Arguni; Endah Supriyati; Gunadi; Hendra Wibawa; Indarto Sulistiyono; Khanza a Vujira; Kristy Iskandar; Ludhang P. Rizki; Marcellus; Mohamad S. Hakim; Nungki Anggorowati; Pramesti G Dewi; Siswanto; Titik Nuryastuti; Tri Wibawa

EPI_ISL_2954684	DKK Gunungkidul/ Puskesmas Girisubo	Genetics Working Group (Pokja Genetik) Faculty of Medicine, Public Health and Nursing Universitas Gadjah Mada (FK-KMK UGM); Disease

Investigation Center Wates Ministry of Agriculture Indonesia; Department of Microbiology FK-KMK UGM; Laboratorium Diagnostik Yayasan Tahija World Mosquito Program (WMP) Yogyakarta Center for Tropical Medicine FK-KMK UGM; Integrated Research Center FK-KMK UGM; Department of Computer Science and Electronics FMIPA UGM; RSUP Dr. Sardjito

Afiahayati; Alvina A Setiawan; Cita S Amalia; Dwi AA Nugrahaningsih; Dwi Indaryati; Edwin W. Daniwijaya; Eggi Arguni; Endah Supriyati; Gunadi; Hendra Wibawa; Indarto Sulistiyono; Kristy Iskandar; Ludhang P. Rizki; Marcellus; Mohamad S. Hakim; Nungki Anggorowati; Pramesti G Dewi; Siswanto; Titik Nuryastuti; Tri Wibawa

EPI_ISL_2955335, EPI_ISL_2955586, EPI_ISL_2955597, EPI_ISL_2955598

DKK Gunungkidul/ Puskesmas Karangmojo I	Genetics Working Group (Pokja Genetik) Faculty of Medicine, Public Health and Nursing Universitas Gadjah Mada (FK-KMK UGM); Disease Investigation Center Wates Ministry of Agriculture Indonesia; Department of Microbiology FK-KMK UGM; Laboratorium Diagnostik Yayasan Tahija World Mosquito Program (WMP) Yogyakarta Center for Tropical Medicine FK-KMK UGM; Integrated Research Center FK-KMK UGM; Department of

Computer Science and Electronics FMIPA UGM; RSUP Dr. Sardjito

Afiahayati; Alvina A Setiawan; Dwi AA Nugrahaningsih; Dwi Indaryati; Dyah A Puspitarani; Edwin W. Daniwijaya; Eggi Arguni; Endah Supriyati; Gunadi; Hana F Hanifin; Hendra Wibawa; Indarto Sulistiyono; Irene Tania; Khanza a Vujira; Kristy Iskandar; Ludhang P. Rizki; Marcellus; Mohamad S. Hakim; Nungki Anggorowati; Pramesti G Dewi; Siswanto; Susan Simanjaya; Titik Nuryastuti; Tri Wibawa

EPI_ISL_2933110	DKK Sleman/Puskesmas Depok 1	Genetics Working Group (Pokja Genetik) Faculty of Medicine, Public Health and Nursing Universitas Gadjah Mada (FK-KMK UGM); Disease

Investigation Center Wates Ministry of Agriculture Indonesia; Department of Microbiology FK-KMK UGM; Laboratorium Diagnostik Yayasan Tahija World Mosquito Program (WMP) Yogyakarta Center for Tropical Medicine FK-KMK UGM; Integrated Research Center FK-KMK UGM; Department of Computer Science and

Afiahayati; Alvina A Setiawan; Dwi AA Nugrahaningsih; Dwi Indaryati; Edwin W. Daniwijaya; Eggi Arguni; Endah Supriyati; Gunadi; Hendra Wibawa; Indarto Sulistiyono; Khanza a Vujira; Kristy Iskandar; Ludhang P. Rizki; Marcellus; Mohamad S. Hakim; Nungki Anggorowati; Pramesti G Dewi; Siswanto; Titik Nuryastuti; Tri Wibawa

EPI_ISL_2932613, EPI_ISL_2933109, EPI_ISL_2943237, EPI_ISL_2955609

EPI_ISL_2955627, EPI_ISL_2955628

DKK Sleman/Puskesmas Depok 1	Genetics Working Group (Pokja Genetik) Faculty of Medicine, Public Health and Nursing Universitas Gadjah Mada (FK-KMK UGM); Disease Investigation Center Wates Ministry of Agriculture Indonesia; Department of Microbiology FK-KMK UGM; Laboratorium Diagnostik Yayasan Tahija World Mosquito Program (WMP) Yogyakarta Center for Tropical Medicine FK-KMK UGM; Integrated Research Center FK-KMK UGM; Department of

Computer Science and Electronics FMIPA UGM; RSUP Dr. Sardjito

DKK Sleman/Puskesmas Ngemplak 2	Genetics Working Group (Pokja Genetik) Faculty of Medicine, Public Health and Nursing Universitas Gadjah Mada (FK-KMK UGM); Disease Investigation Center Wates Ministry of Agriculture Indonesia; Department of Microbiology FK-KMK UGM; Laboratorium Diagnostik Yayasan Tahija World Mosquito Program (WMP) Yogyakarta Center for Tropical Medicine FK-KMK UGM; Integrated Research Center FK-KMK UGM; Department of

Computer Science and Electronics FMIPA UGM; RSUP Dr. Sardjito

Afiahayati; Alvina A Setiawan; Cita S Amalia; Dwi AA Nugrahaningsih; Dwi Indaryati; Dyah A Puspitarani; Edwin W. Daniwijaya; Eggi Arguni; Endah Supriyati; Gunadi; Hana F Hanifin; Hendra Wibawa; Indarto Sulistiyono; Irene Tania; Kristy Iskandar; Ludhang P. Rizki; Marcellus; Mohamad S. Hakim; Nungki Anggorowati; Pramesti G Dewi; Siswanto; Susan Simanjaya; Titik Nuryastuti; Tri Wibawa

Afiahayati; Dwi AA Nugrahaningsih; Dwi Indaryati; Dyah A Puspitarani; Edwin W. Daniwijaya; Eggi Arguni; Endah Supriyati; Gunadi; Hana F Hanifin; Hendra Wibawa; Indarto Sulistiyono; Irene Tania; Kristy Iskandar; Ludhang P. Rizki; Marcellus; Mohamad S. Hakim; Nungki Anggorowati; Pramesti G Dewi; Siswanto; Susan Simanjaya; Titik Nuryastuti; Tri Wibawa

EPI_ISL_2932610	KOTA Yogya DKK Gondokusuman II	Genetics Working Group (Pokja Genetik) Faculty of Medicine, Public Health and Nursing Universitas Gadjah Mada (FK-KMK UGM); Disease

Investigation Center Wates Ministry of Agriculture Indonesia; Department of Microbiology FK-KMK UGM; Laboratorium Diagnostik Yayasan Tahija World Mosquito Program (WMP) Yogyakarta Center for

EPI_ISL_2964681	Puskesmas Colomadu 1 DKK Karanganyar	Genetics Working Group (Pokja Genetik) Faculty of Medicine, Public Health and Nursing Universitas Gadjah Mada (FK-KMK UGM); Disease

Investigation Center Wates Ministry of Agriculture Indonesia; Department of Microbiology FK-KMK UGM; Laboratorium Diagnostik Yayasan Tahija World Mosquito Program (WMP) Yogyakarta Center for Tropical Medicine FK-KMK UGM; Integrated Research Center FK-KMK UGM; Department of Computer Science and Electronics FMIPA UGM; RSUP Dr. Sardjito

Afiahayati; Alvina A Setiawan; Dwi AA Nugrahaningsih; Dwi Indaryati; Edwin W. Daniwijaya; Eggi Arguni; Endah Supriyati; Gunadi; Hendra Wibawa; Indarto Sulistiyono; Khanza a Vujira; Kristy Iskandar; Ludhang P. Rizki; Marcellus; Mohamad S. Hakim; Nungki Anggorowati; Pramesti G Dewi; Siswanto; Titik Nuryastuti; Tri Wibawa

Afiahayati; Alvina A Setiawan; Dwi AA Nugrahaningsih; Dwi Indaryati; Edwin W. Daniwijaya; Eggi Arguni; Endah Supriyati; Gunadi; Hendra Wibawa; Indarto Sulistiyono; Khanza a Vujira; Kristy Iskandar; Ludhang P. Rizki; Marcellus; Mohamad S. Hakim; Nungki Anggorowati; Pramesti G Dewi; Siswanto; Titik Nuryastuti; Tri Wibawa

EPI_ISL_2964917, EPI_ISL_2964918, EPI_ISL_2964919, EPI_ISL_2964920, EPI_ISL_2964921

EPI_ISL_2964707, EPI_ISL_2964872

RS Dr. OEN KANDANG SAPI SOLO	Genetics Working Group (Pokja Genetik) Faculty of Medicine, Public Health and Nursing Universitas Gadjah Mada (FK-KMK UGM); Disease Investigation Center Wates Ministry of Agriculture Indonesia; Department of Microbiology FK-KMK UGM; Laboratorium Diagnostik Yayasan Tahija World Mosquito Program (WMP) Yogyakarta Center for Tropical Medicine FK-KMK UGM; Integrated Research Center FK-KMK UGM; Department of

Computer Science and Electronics FMIPA UGM; RSUP Dr. Sardjito

RS Dr. Oen Kandang Sapi Solo	Genetics Working Group (Pokja Genetik) Faculty of Medicine, Public Health and Nursing Universitas Gadjah Mada (FK-KMK UGM); Disease Investigation Center Wates Ministry of Agriculture Indonesia; Department of Microbiology FK-KMK UGM; Laboratorium Diagnostik Yayasan Tahija World Mosquito Program (WMP) Yogyakarta Center for Tropical Medicine FK-KMK UGM; Integrated Research Center FK-KMK UGM; Department of

Computer Science and Electronics FMIPA UGM; RSUP Dr. Sardjito

Afiahayati; Alvina A Setiawan; Cita S Amalia; Dwi AA Nugrahaningsih; Dwi Indaryati; Dyah A Puspitarani; Edwin W. Daniwijaya; Eggi Arguni; Endah Supriyati; Gunadi; Hana F Hanifin; Hendra Wibawa; Indarto Sulistiyono; Irene Tania; Khanza a Vujira; Kristy Iskandar; Ludhang P. Rizki; Marcellus; Mohamad S. Hakim; Nungki Anggorowati; Pramesti G Dewi; Siswanto; Susan Simanjaya; Titik Nuryastuti; Tri Wibawa

Afiahayati; Alvina A Setiawan; Cita S Amalia; Dwi AA Nugrahaningsih; Dwi Indaryati; Dyah A Puspitarani; Edwin W. Daniwijaya; Eggi Arguni; Endah Supriyati; Gunadi; Hendra Wibawa; Indarto Sulistiyono; Kristy Iskandar; Ludhang P. Rizki; Marcellus; Mohamad S. Hakim; Nungki Anggorowati; Pramesti G Dewi; Siswanto; Susan Simanjaya; Titik Nuryastuti; Tri Wibawa

EPI_ISL_2955608	RSA UGM	Genetics Working Group (Pokja Genetik) Faculty of Medicine, Public Health and Nursing Universitas Gadjah Mada (FK-KMK UGM); Disease

Investigation Center Wates Ministry of Agriculture Indonesia; Department of Microbiology FK-KMK UGM; Laboratorium Diagnostik Yayasan Tahija World Mosquito Program (WMP) Yogyakarta Center for Tropical Medicine FK-KMK UGM; Integrated Research Center FK-KMK UGM; Department of Computer Science and Electronics FMIPA UGM; RSUP Dr. Sardjito

EPI_ISL_2964684, EPI_ISL_2964688, EPI_ISL_2964692, EPI_ISL_2964941, EPI_ISL_2964942, EPI_ISL_2964943, EPI_ISL_2964944, EPI_ISL_2964956, EPI_ISL_2964957

see above	RSUD DR. MOEWARDI	Genetics Working Group (Pokja Genetik) Faculty of Medicine, Public Health and Nursing Universitas Gadjah Mada (FK-KMK UGM); Disease

Investigation Center Wates Ministry of Agriculture Indonesia; Department of Microbiology FK-KMK UGM; Laboratorium Diagnostik Yayasan Tahija World Mosquito Program (WMP) Yogyakarta Center for Tropical Medicine FK-KMK UGM; Integrated Research Center FK-KMK UGM; Department of Computer Science and Electronics FMIPA UGM; RSUP Dr. Sardjito

EPI_ISL_2964677	RSUD DR. MOEWARDI Surakarta	Genetics Working Group (Pokja Genetik) Faculty of Medicine, Public Health and Nursing Universitas Gadjah Mada (FK-KMK UGM); Disease

Investigation Center Wates Ministry of Agriculture Indonesia; Department of Microbiology FK-KMK UGM; Laboratorium Diagnostik Yayasan Tahija World Mosquito Program (WMP) Yogyakarta Center for Tropical Medicine FK-KMK UGM; Integrated Research Center FK-KMK UGM; Department of Computer Science and Electronics FMIPA UGM; RSUP Dr. Sardjito

Afiahayati; Dwi AA Nugrahaningsih; Dwi Indaryati; Dyah A Puspitarani; Edwin W. Daniwijaya; Eggi Arguni; Endah Supriyati; Gunadi; Hendra Wibawa; Indarto Sulistiyono; Kristy Iskandar; Ludhang P. Rizki; Marcellus; Mohamad S. Hakim; Nungki Anggorowati; Pramesti G Dewi; Siswanto; Susan Simanjaya; Titik Nuryastuti; Tri Wibawa

Afiahayati; Alvina A Setiawan; Cita S Amalia; Dwi AA Nugrahaningsih; Dwi Indaryati; Dyah A Puspitarani; Edwin W. Daniwijaya; Eggi Arguni; Endah Supriyati; Gunadi; Hana F Hanifin; Hendra Wibawa; I Putu Aditio A; Indarto Sulistiyono; Irene Tania; Kristy Iskandar; Ludhang P. Rizki; Marcellus; Mohamad S. Hakim; Nungki Anggorowati; Pramesti G Dewi; Siswanto; Susan Simanjaya; Titik Nuryastuti; Tri Wibawa

Afiahayati; Alvina A Setiawan; Dwi AA Nugrahaningsih; Dwi Indaryati; Edwin W. Daniwijaya; Eggi Arguni; Endah Supriyati; Gunadi; Hendra Wibawa; Indarto Sulistiyono; Khanza a Vujira; Kristy Iskandar; Ludhang P. Rizki; Marcellus; Mohamad S. Hakim; Nungki Anggorowati; Pramesti G Dewi; Siswanto; Titik Nuryastuti; Tri Wibawa

EPI_ISL_2954064	RSUD Panembahan Senopati, Bantul	Genetics Working Group (Pokja Genetik) Faculty of Medicine, Public Health and Nursing Universitas Gadjah Mada (FK-KMK UGM); Disease

Investigation Center Wates Ministry of Agriculture Indonesia; Department of Microbiology FK-KMK UGM; Laboratorium Diagnostik Yayasan Tahija World Mosquito Program (WMP) Yogyakarta Center for Tropical Medicine FK-KMK UGM; Integrated Research Center FK-KMK UGM; Department of Computer

Afiahayati; Dwi AA Nugrahaningsih; Dwi Indaryati; Edwin W. Daniwijaya; Eggi Arguni; Endah Supriyati; Gunadi; Hana F Hanifin; Hendra Wibawa; Indarto Sulistiyono; Irene Tania; Kristy Iskandar; Ludhang P. Rizki; Marcellus; Mohamad S. Hakim; Nungki Anggorowati; Pramesti G Dewi; Siswanto; Titik Nuryastuti; Tri Wibawa

EPI_ISL_2502616, EPI_ISL_2502627, EPI_ISL_2502628, EPI_ISL_2502629, EPI_ISL_2502667, EPI_ISL_2502687, EPI_ISL_2502717, EPI_ISL_2502763, EPI_ISL_2502764, EPI_ISL_2502765, EPI_ISL_2506039, EPI_ISL_2506040, EPI_ISL_2506041, EPI_ISL_2534078, EPI_ISL_2534079, EPI_ISL_2534082, EPI_ISL_2534289, EPI_ISL_2534338, EPI_ISL_2534346, EPI_ISL_2534376, EPI_ISL_2534377, EPI_ISL_2534379, EPI_ISL_2534454, EPI_ISL_2534456, EPI_ISL_2534457, EPI_ISL_2534458, EPI_ISL_2534490, EPI_ISL_2534518

see above	RSUD dr. LOEKMONO HADI	Genetics Working Group (Pokja Genetik) Faculty of Medicine, Public Health and Nursing Universitas Gadjah Mada (FK-KMK UGM); Disease

Investigation Center Wates Ministry of Agriculture Indonesia; Department of Microbiology FK-KMK UGM; Laboratorium Diagnostik Yayasan Tahija World Mosquito Program (WMP) Yogyakarta Center for Tropical Medicine FK-KMK UGM; Integrated Research Center FK-KMK UGM; Department of Computer Science and Electronics FMIPA UGM; RSUP Dr. Sardjito

EPI_ISL_2955607	RSUP Dr Sardjito	Genetics Working Group (Pokja Genetik) Faculty of Medicine, Public Health and Nursing Universitas Gadjah Mada (FK-KMK UGM); Disease

Investigation Center Wates Ministry of Agriculture Indonesia; Department of Microbiology FK-KMK UGM; Laboratorium Diagnostik Yayasan Tahija World Mosquito Program (WMP) Yogyakarta Center for Tropical Medicine FK-KMK UGM; Integrated Research Center FK-KMK UGM; Department of Computer Science and Electronics FMIPA UGM; RSUP Dr. Sardjito

EPI_ISL_2943173	RSUP Dr. Sardjito	Genetics Working Group (Pokja Genetik) Faculty of Medicine, Public Health and Nursing Universitas Gadjah Mada (FK-KMK UGM); Disease

Investigation Center Wates Ministry of Agriculture Indonesia; Department of Microbiology FK-KMK UGM; Laboratorium Diagnostik Yayasan Tahija World Mosquito Program

EPI_ISL_2955606	RSUP Sardjito	Genetics Working Group (Pokja Genetik) Faculty of Medicine, Public Health and Nursing Universitas Gadjah Mada (FK-KMK UGM); Disease

Investigation Center Wates Ministry of Agriculture Indonesia; Department of Microbiology FK-KMK UGM; Laboratorium Diagnostik Yayasan Tahija World Mosquito Program (WMP) Yogyakarta Center for Tropical Medicine FK-KMK UGM; Integrated Research Center FK-KMK UGM; Department of Computer Science and Electronics FMIPA UGM; RSUP Dr. Sardjito

Afiahayati; Alvina A Setiawan; Cita S Amalia; Dwi AA Nugrahaningsih; Dwi Indaryati; Dyah A Puspitarani; Edwin W. Daniwijaya; Eggi Arguni; Endah Supriyati; Gunadi; Hana F Hanifin; Hendra Wibawa; Indarto Sulistiyono; Irene Tania; Khanza a Vujira; Kristy Iskandar; Ludhang P. Rizki; Marcellus; Mohamad S. Hakim; Nungki Anggorowati; Pramesti G Dewi; Siswanto; Susan Simanjaya; Titik Nuryastuti; Tri Wibawa

. Marcellus; Afiahayati; Bambang Sigit Riyanto; Dwi AA Nugrahaningsih; Edwin W. Daniwijaya; Eggi Arguni; Eko Budiono; Endah Supriyati; Gunadi; Hana F Hanifin; Hendra Wibawa; Heni Retnowulan; Ika Trisnawati; Ira Puspitawati; Irene Tania; Kristy Iskandar; Ludhang P. Rizki; Mohamad S. Hakim; Munawar Gani; Nungki Anggorowati; Nur Imma Fatimah Harahap; Nur Rahmi Ananda; Osman Sianipar; Riat El Khair; Satria Maulana; Siswanto; Sumardi; Titik Nuryastuti; Tri Wibawa; Umi Solekhah Intansari; Yunika Puspadewi; Elizabeth Henny Herningtiyas

. Marcellus; Afiahayati; Bambang Sigit Riyanto; Dwi AA Nugrahaningsih; Edwin W. Daniwijaya; Eggi Arguni; Eko Budiono; Endah Supriyati; Gunadi; Hana F Hanifin; Hendra Wibawa; Heni Retnowulan; Ika Trisnawati; Ira Puspitawati; Irene Tania; Kristy Iskandar; Ludhang P. Rizki; Mohamad S. Hakim; Munawar Gani; Nungki Anggorowati; Nur Imma Fatimah Harahap; Nur Rahmi Ananda; Osman Sianipar; Riat El Khair; Satria Maulana; Siswanto; Sumardi; Titik Nuryastuti; Tri Wibawa; Umi Solekhah Intansari; Yunika Puspadewi; Elizabeth Henny Herningtiyas

. Marcellus; Afiahayati; Alvina A Setiawan; Bambang Sigit Riyanto; Dwi AA Nugrahaningsih; Edwin W. Daniwijaya; Eggi Arguni; Eko Budiono; Endah Supriyati; Gunadi; Hendra Wibawa; Heni Retnowulan; Ika Trisnawati; Ira Puspitawati; Khanza a Vujira; Kristy Iskandar; Ludhang P. Rizki; Mohamad S. Hakim; Munawar Gani; Nungki Anggorowati; Nur Imma Fatimah Harahap; Nur Rahmi Ananda; Osman Sianipar; Riat El Khair; Satria Maulana; Siswanto; Sumardi; Titik Nuryastuti; Tri Wibawa; Umi Solekhah Intansari; Yunika Puspadewi; Elizabeth Henny Herningtiyas

### Acknowledgement EPI_SET Identifier: EPI_SET_20220426dx

#### Accession ID Originating Laboratory Submitting Laboratory Authors

EPI_ISL_9433340, EPI_ISL_9433342, EPI_ISL_9433345, EPI_ISL_9433347, EPI_ISL_9433399, EPI_ISL_9487459

EPI_ISL_9433338, EPI_ISL_9487460

EPI_ISL_9435593, EPI_ISL_9435799

BB VET Wates/Puskesmas Kalibawang Wates	Genetics Working Group (Pokja Genetik) Faculty of Medicine, Public Health and Nursing Universitas Gadjah Mada (FK-KMK UGM);

Disease Investigation Center Wates Ministry of Agriculture Indonesia; Department of Microbiology FK-KMK UGM; Laboratorium Diagnostik Yayasan Tahija World Mosquito Program (WMP) Yogyakarta Center for Tropical Medicine FK-KMK UGM; Integrated Research Center FK-KMK UGM; Department of Computer Science and Electronics FMIPA UGM; RSUP Dr. Sardjito

BB VET Wates/Puskesmas Samigaluh I Wates	Genetics Working Group (Pokja Genetik) Faculty of Medicine, Public Health and Nursing Universitas Gadjah Mada (FK-KMK UGM);

Disease Investigation Center Wates Ministry of Agriculture Indonesia; Department of Microbiology FK-KMK UGM; Laboratorium Diagnostik Yayasan Tahija World Mosquito Program (WMP) Yogyakarta Center for Tropical Medicine FK-KMK UGM; Integrated Research Center FK-KMK UGM; Department of Computer Science and Electronics FMIPA UGM; RSUP Dr. Sardjito

BB VET Wates/Puskesmas Wates	Genetics Working Group (Pokja Genetik) Faculty of Medicine, Public Health and Nursing Universitas Gadjah Mada (FK-KMK UGM); Disease Investigation Center Wates Ministry of Agriculture Indonesia; Department of Microbiology FK-KMK UGM; Laboratorium Diagnostik Yayasan Tahija World Mosquito Program (WMP) Yogyakarta Center for Tropical Medicine FK-KMK UGM; Integrated

Research Center FK-KMK UGM; Department of Computer Science and Electronics FMIPA UGM; RSUP Dr. Sardjito

Afiahayati; Dwi AA Nugrahaningsih; Dwi Indaryati; Dyah A Puspitarani; Edwin W. Daniwijaya; Eggi Arguni; Endah Supriyati; Fadila D T Utami; Gita C Gabriela; Gunadi; Hendra Wibawa; Ika Trisnawati; Indarto Sulistiyono; Khanza A Vujira; Kristy Iskandar; Lanang Aditama; Laudria S Eryvinka; Ludhang P. Rizki; Mohamad S. Hakim; Nungki Anggorowati; Pramesti G Dewi; Riat E Khair; Siswanto; Titik Nuryastuti; Tri Wibawa

Afiahayati; Dwi AA Nugrahaningsih; Dwi Indaryati; Dyah A Puspitarani; Edwin W. Daniwijaya; Eggi Arguni; Endah Supriyati; Fadila D T Utami; Gita C Gabriela; Gunadi; Hendra Wibawa; Ika Trisnawati; Indarto Sulistiyono; Khanza A Vujira; Kristy Iskandar; Lanang Aditama; Laudria S Eryvinka; Ludhang P. Rizki; Mohamad S. Hakim; Nungki Anggorowati; Pramesti G Dewi; Riat E Khair; Siswanto; Titik Nuryastuti; Tri Wibawa

Afiahayati; Dwi AA Nugrahaningsih; Dwi Indaryati; Dyah A Puspitarani; Edwin W. Daniwijaya; Eggi Arguni; Endah Supriyati; Gita C Gabriela; Gunadi; Hendra Wibawa; Indarto Sulistiyono; Khanza A Vujira; Kristy Iskandar; Ludhang P. Rizki; Mohamad S. Hakim; Nungki Anggorowati; Pramesti G Dewi; Siswanto; Titik Nuryastuti; Tri Wibawa

EPI_ISL_12241047	BBTKLPP	Genetics Working Group (Pokja Genetik) Faculty of Medicine, Public Health and Nursing Universitas Gadjah Mada (FK-KMK UGM);

Disease Investigation Center Wates Ministry of Agriculture Indonesia; Department of Microbiology FK-KMK UGM; Laboratorium Diagnostik Yayasan Tahija World Mosquito Program (WMP) Yogyakarta Center for Tropical Medicine FK-KMK UGM; Integrated Research Center FK-KMK UGM; Department of Computer Science and Electronics FMIPA UGM; RSUP Dr. Sardjito

EPI_ISL_9487466, EPI_ISL_9487490, EPI_ISL_9487491, EPI_ISL_9570708, EPI_ISL_9570710, EPI_ISL_9570712, EPI_ISL_9570714, EPI_ISL_9570715, EPI_ISL_9570717, EPI_ISL_9570720, EPI_ISL_9570721, EPI_ISL_9570722

see above	Balitbangkes	Genetics Working Group (Pokja Genetik) Faculty of Medicine, Public Health and Nursing Universitas Gadjah Mada (FK-KMK UGM);

Disease Investigation Center Wates Ministry of Agriculture Indonesia; Department of Microbiology FK-KMK UGM; Laboratorium Diagnostik Yayasan Tahija World Mosquito Program (WMP) Yogyakarta Center for Tropical Medicine FK-KMK UGM; Integrated Research Center FK-KMK UGM; Department of Computer Science and Electronics FMIPA UGM; RSUP Dr. Sardjito

EPI_ISL_9433333	DKK Yogyakarta	Genetics Working Group (Pokja Genetik) Faculty of Medicine, Public Health and Nursing Universitas Gadjah Mada (FK-KMK UGM);

Disease Investigation Center Wates Ministry of Agriculture Indonesia; Department of Microbiology FK-KMK UGM; Laboratorium Diagnostik Yayasan Tahija World Mosquito Program (WMP) Yogyakarta Center for Tropical Medicine FK-KMK UGM; Integrated Research Center FK-KMK UGM; Department of Computer Science and Electronics FMIPA UGM; RSUP Dr. Sardjito

Afiahayati; Dwi AA Nugrahaningsih; Dyah A Puspitarani; Edita M Devana; Edwin W. Daniwijaya; Eggi Arguni; Endah Supriyati; Esensi T Geometri; Fadila D T Utami; Gita C Gabriela; Gunadi; Hendra Wibawa; Ika Trisnawati; Khanza A Vujira; Kristy Iskandar; Ludhang P. Rizki; Mohamad S. Hakim; Nungki Anggorowati; Riat E Khair; Siswanto; Titik Nuryastuti; Tri Wibawa; Verrell Christopher

Afiahayati; Dwi AA Nugrahaningsih; Dyah A Puspitarani; Edwin W. Daniwijaya; Eggi Arguni; Endah Supriyati; Fadila D T Utami; Gita C Gabriela; Gunadi; Hendra Wibawa; Ika Trisnawati; Khanza A Vujira; Kristy Iskandar; Lanang Aditama; Laudria S Eryvinka; Ludhang P. Rizki; Mohamad S. Hakim; Nungki Anggorowati; Riat E Khair; Siswanto; Titik Nuryastuti; Tri Wibawa

Afiahayati; Dwi AA Nugrahaningsih; Dwi Indaryati; Dyah A Puspitarani; Edwin W. Daniwijaya; Eggi Arguni; Endah Supriyati; Gita Christy Gabriela; Gunadi; Hendra Wibawa; Indarto Sulistiyono; Khanza A Vujira; Kristy Iskandar; Ludhang P. Rizki; Mohamad S. Hakim; Nungki Anggorowati; Pramesti G Dewi; Siswanto; Titik Nuryastuti; Tri Wibawa

EPI_ISL_9487451, EPI_ISL_9487452, EPI_ISL_9487453, EPI_ISL_9487454

EPI_ISL_7162207, EPI_ISL_7162210

DKK Yogyakarta/HI-LAB Yogyakarta	Genetics Working Group (Pokja Genetik) Faculty of Medicine, Public Health and Nursing Universitas Gadjah Mada (FK-KMK UGM); Disease Investigation Center Wates Ministry of Agriculture Indonesia; Department of Microbiology FK-KMK UGM; Laboratorium Diagnostik Yayasan Tahija World Mosquito Program (WMP) Yogyakarta Center for Tropical Medicine FK-KMK UGM; Integrated

Research Center FK-KMK UGM; Department of Computer Science and Electronics FMIPA UGM; RSUP Dr. Sardjito

Dinkes Kab Demak	Genetics Working Group (Pokja Genetik) Faculty of Medicine, Public Health and Nursing Universitas Gadjah Mada (FK-KMK UGM); Disease Investigation Center Wates Ministry of Agriculture Indonesia; Department of Microbiology FK-KMK UGM; Laboratorium Diagnostik Yayasan Tahija World Mosquito Program (WMP) Yogyakarta Center for Tropical Medicine FK-KMK UGM; Integrated

Research Center FK-KMK UGM; Department of Computer Science and Electronics FMIPA UGM; RSUP Dr. Sardjito

Afiahayati; Dwi AA Nugrahaningsih; Dyah A Puspitarani; Edwin W. Daniwijaya; Eggi Arguni; Endah Supriyati; Fadila D T Utami; Gita C Gabriela; Gunadi; Hendra Wibawa; Ika Trisnawati; Khanza A Vujira; Kristy Iskandar; Lanang Aditama; Laudria S Eryvinka; Ludhang P. Rizki; Mohamad S. Hakim; Nungki Anggorowati; Riat E Khair; Siswanto; Titik Nuryastuti; Tri Wibawa

Afiahayati; Dwi AA Nugrahaningsih; Dwi Indaryati; Dyah A Puspitarani; Edwin W. Daniwijaya; Eggi Arguni; Endah Supriyati; Gita C Gabriela; Gunadi; Hendra Wibawa; Indarto Sulistiyono; Khanza A Vujira; Kristy Iskandar; Ludhang P. Rizki; Marcellus; Mohamad S. Hakim; Nungki Anggorowati; Pramesti G Dewi; Siswanto; Titik Nuryastuti; Tri Wibawa

EPI_ISL_10658791	Dr. Oen Hospital	Genetics Working Group (Pokja Genetik) Faculty of Medicine, Public Health and Nursing Universitas Gadjah Mada (FK-KMK UGM);

Disease Investigation Center Wates Ministry of Agriculture Indonesia; Department of Microbiology FK-KMK UGM; Laboratorium Diagnostik Yayasan Tahija World Mosquito Program (WMP) Yogyakarta Center for Tropical Medicine FK-KMK UGM; Integrated Research Center FK-KMK UGM; Department of Computer Science and Electronics FMIPA UGM; RSUP Dr. Sardjito

Afiahayati; Dwi AA Nugrahaningsih; Dyah A Puspitarani; Edita M Devana; Edwin W. Daniwijaya; Eggi Arguni; Endah Supriyati; Esensi T Geometri; Fadila D T Utami; Gita C Gabriela; Gunadi; Hendra Wibawa; Ika Trisnawati; Khanza A Vujira; Kristy Iskandar; Ludhang P. Rizki; Mohamad S. Hakim; Nungki Anggorowati; Riat E Khair; Siswanto; Titik Nuryastuti; Tri Wibawa; Verrell Christopher

EPI_ISL_10657578, EPI_ISL_10657730, EPI_ISL_10658771

EPI_ISL_10657580, EPI_ISL_10657732

EPI_ISL_10639786, EPI_ISL_10657735, EPI_ISL_10658762, EPI_ISL_10658767, EPI_ISL_10658768

Dr. Oen Solo Baru Hospital	Genetics Working Group (Pokja Genetik) Faculty of Medicine, Public Health and Nursing Universitas Gadjah Mada (FK-KMK UGM); Disease Investigation Center Wates Ministry of Agriculture Indonesia; Department of Microbiology FK-KMK UGM; Laboratorium Diagnostik Yayasan Tahija World Mosquito Program (WMP) Yogyakarta Center for Tropical Medicine FK-KMK UGM; Integrated

Research Center FK-KMK UGM; Department of Computer Science and Electronics FMIPA UGM; RSUP Dr. Sardjito

Hospital District. Wonosobo	Genetics Working Group (Pokja Genetik) Faculty of Medicine, Public Health and Nursing Universitas Gadjah Mada (FK-KMK UGM); Disease Investigation Center Wates Ministry of Agriculture Indonesia; Department of Microbiology FK-KMK UGM; Laboratorium Diagnostik Yayasan Tahija World Mosquito Program (WMP) Yogyakarta Center for Tropical Medicine FK-KMK UGM; Integrated

Research Center FK-KMK UGM; Department of Computer Science and Electronics FMIPA UGM; RSUP Dr. Sardjito

JIH SOLO	Genetics Working Group (Pokja Genetik) Faculty of Medicine, Public Health and Nursing Universitas Gadjah Mada (FK-KMK UGM); Disease Investigation Center Wates Ministry of Agriculture Indonesia; Department of Microbiology FK-KMK UGM; Laboratorium Diagnostik Yayasan Tahija World Mosquito Program (WMP) Yogyakarta Center for Tropical Medicine FK-KMK UGM; Integrated

Research Center FK-KMK UGM; Department of Computer Science and Electronics FMIPA UGM; RSUP Dr. Sardjito

Afiahayati; Dwi AA Nugrahaningsih; Dyah A Puspitarani; Edita M Devana; Edwin W. Daniwijaya; Eggi Arguni; Endah Supriyati; Esensi T Geometri; Fadila D T Utami; Gita C Gabriela; Gunadi; Hendra Wibawa; Ika Trisnawati; Khanza A Vujira; Kristy Iskandar; Ludhang P. Rizki; Mohamad S. Hakim; Nungki Anggorowati; Riat E Khair; Siswanto; Titik Nuryastuti; Tri Wibawa; Verrell Christopher

Afiahayati; Dwi AA Nugrahaningsih; Dyah A Puspitarani; Edita M Devana; Edwin W. Daniwijaya; Eggi Arguni; Endah Supriyati; Esensi T Geometri; Fadila D T Utami; Gita C Gabriela; Gunadi; Hendra Wibawa; Ika Trisnawati; Khanza A Vujira; Kristy Iskandar; Ludhang P. Rizki; Mohamad S. Hakim; Nungki Anggorowati; Riat E Khair; Siswanto; Titik Nuryastuti; Tri Wibawa; Verrell Christopher

Afiahayati; Dwi AA Nugrahaningsih; Dyah A Puspitarani; Edita M Devana; Edwin W. Daniwijaya; Eggi Arguni; Endah Supriyati; Esensi T Geometri; Fadila D T Utami; Gita C Gabriela; Gunadi; Hendra Wibawa; Ika Trisnawati; Khanza A Vujira; Kristy Iskandar; Ludhang P. Rizki; Mohamad S. Hakim; Nungki Anggorowati; Riat E Khair; Siswanto; Titik Nuryastuti; Tri Wibawa; Verrell Christopher

EPI_ISL_9702359, EPI_ISL_9702419, EPI_ISL_9702573, EPI_ISL_9702602, EPI_ISL_9702615, EPI_ISL_9713699, EPI_ISL_9713708, EPI_ISL_9713709, EPI_ISL_9713710, EPI_ISL_9713711, EPI_ISL_9713712, EPI_ISL_9753879, EPI_ISL_9754267, EPI_ISL_9754269, EPI_ISL_9754270, EPI_ISL_12241131, EPI_ISL_12241581

see above	Klinik Cito DIY	Genetics Working Group (Pokja Genetik) Faculty of Medicine, Public Health and Nursing Universitas Gadjah Mada (FK-KMK UGM);

Disease Investigation Center Wates Ministry of Agriculture Indonesia; Department of Microbiology FK-KMK UGM; Laboratorium Diagnostik Yayasan Tahija World Mosquito Program (WMP) Yogyakarta Center for Tropical Medicine FK-KMK UGM; Integrated Research Center FK-KMK UGM; Department of Computer Science and Electronics FMIPA UGM; RSUP Dr. Sardjito

Afiahayati; Dwi AA Nugrahaningsih; Dyah A Puspitarani; Edwin W. Daniwijaya; Eggi Arguni; Endah Supriyati; Fadila D T Utami; Gita C Gabriela; Gunadi; Hendra Wibawa; Ika Trisnawati; Khanza A Vujira; Kristy Iskandar; Lanang Aditama; Laudria S Eryvinka; Ludhang P. Rizki; Mohamad S. Hakim; Nungki Anggorowati; Riat E Khair; Siswanto; Titik Nuryastuti; Tri Wibawa

EPI_ISL_7160934, EPI_ISL_7160951, EPI_ISL_7160953, EPI_ISL_7160957, EPI_ISL_7162054, EPI_ISL_12241065

EPI_ISL_7160420, EPI_ISL_7160930, EPI_ISL_12241067, EPI_ISL_12241068

LAB PCR RSUD CILACAP	Genetics Working Group (Pokja Genetik) Faculty of Medicine, Public Health and Nursing Universitas Gadjah Mada (FK-KMK UGM); Disease Investigation Center Wates Ministry of Agriculture Indonesia; Department of Microbiology FK-KMK UGM; Laboratorium Diagnostik Yayasan Tahija World Mosquito Program (WMP) Yogyakarta Center for Tropical Medicine FK-KMK UGM; Integrated

Research Center FK-KMK UGM; Department of Computer Science and Electronics FMIPA UGM; RSUP Dr. Sardjito

LAB RS Pertamina CILACAP	Genetics Working Group (Pokja Genetik) Faculty of Medicine, Public Health and Nursing Universitas Gadjah Mada (FK-KMK UGM); Disease Investigation Center Wates Ministry of Agriculture Indonesia; Department of Microbiology FK-KMK UGM; Laboratorium Diagnostik Yayasan Tahija World Mosquito Program (WMP) Yogyakarta Center for Tropical Medicine FK-KMK UGM; Integrated

Research Center FK-KMK UGM; Department of Computer Science and Electronics FMIPA UGM; RSUP Dr. Sardjito

. Marcellus; Afiahayati; Alvina A Setiawan; Bambang Sigit Riyanto; Dwi AA Nugrahaningsih; Dwi Indaryati; Dyah A Puspitarani; Edwin W. Daniwijaya; Eggi Arguni; Eko Budiono; Endah Supriyati; Gita C Gabriela; Gunadi; Hendra Wibawa; Heni Retnowulan; Ika Trisnawati; Indarto Sulistiyono; Ira Puspitawati; Khanza A Vujira; Khanza a Vujira; Kristy Iskandar; Ludhang P. Rizki; Marcellus; Mohamad S. Hakim; Munawar Gani; Nungki Anggorowati; Nur Imma Fatimah Harahap; Nur Rahmi Ananda; Osman Sianipar; Pramesti G Dewi; Riat El Khair; Satria Maulana; Siswanto; Sumardi; Susan Simanjaya; Titik Nuryastuti; Tri Wibawa; Umi Solekhah Intansari; Yunika Puspadewi; Elizabeth Henny Herningtiyas

Afiahayati; Dwi AA Nugrahaningsih; Dwi Indaryati; Dyah A Puspitarani; Edwin W. Daniwijaya; Eggi Arguni; Endah Supriyati; Gita C Gabriela; Gunadi; Hendra Wibawa; Indarto Sulistiyono; Khanza A Vujira; Khanza a Vujira; Kristy Iskandar; Ludhang P. Rizki; Marcellus; Mohamad S. Hakim; Nungki Anggorowati; Pramesti G Dewi; Siswanto; Susan Simanjaya; Titik Nuryastuti; Tri Wibawa

EPI_ISL_9754617, EPI_ISL_9754619, EPI_ISL_9754620, EPI_ISL_9754765, EPI_ISL_9754857, EPI_ISL_9754978, EPI_ISL_9755096, EPI_ISL_9755169, EPI_ISL_9755171

see above	Lab Intibios	Genetics Working Group (Pokja Genetik) Faculty of Medicine, Public Health and Nursing Universitas Gadjah Mada (FK-KMK UGM);

Disease Investigation Center Wates Ministry of Agriculture Indonesia; Department of Microbiology FK-KMK UGM; Laboratorium Diagnostik Yayasan Tahija World Mosquito Program (WMP) Yogyakarta Center for Tropical Medicine FK-KMK UGM; Integrated Research Center FK-KMK UGM; Department of Computer Science and Electronics FMIPA UGM; RSUP Dr. Sardjito

Afiahayati; Dwi AA Nugrahaningsih; Dyah A Puspitarani; Edwin W. Daniwijaya; Eggi Arguni; Endah Supriyati; Fadila D T Utami; Gita C Gabriela; Gunadi; Hendra Wibawa; Ika Trisnawati; Khanza A Vujira; Kristy Iskandar; Lanang Aditama; Laudria S Eryvinka; Ludhang P. Rizki; Mohamad S. Hakim; Nungki Anggorowati; Riat E Khair; Siswanto; Titik Nuryastuti; Tri Wibawa

EPI_ISL_9831384, EPI_ISL_9831597, EPI_ISL_9831736

EPI_ISL_9434160, EPI_ISL_9487412

Lab Microbiology, FK-KMK UGM	Genetics Working Group (Pokja Genetik) Faculty of Medicine, Public Health and Nursing Universitas Gadjah Mada (FK-KMK UGM); Disease Investigation Center Wates Ministry of Agriculture Indonesia; Department of Microbiology FK-KMK UGM; Laboratorium Diagnostik Yayasan Tahija World Mosquito Program (WMP) Yogyakarta Center for Tropical Medicine FK-KMK UGM; Integrated

Research Center FK-KMK UGM; Department of Computer Science and Electronics FMIPA UGM; RSUP Dr. Sardjito

Lab RS Bethesda	Genetics Working Group (Pokja Genetik) Faculty of Medicine, Public Health and Nursing Universitas Gadjah Mada (FK-KMK UGM); Disease Investigation Center Wates Ministry of Agriculture Indonesia; Department of Microbiology FK-KMK UGM; Laboratorium Diagnostik Yayasan Tahija World Mosquito Program (WMP) Yogyakarta Center for Tropical Medicine FK-KMK UGM; Integrated

Research Center FK-KMK UGM; Department of Computer Science and Electronics FMIPA UGM; RSUP Dr. Sardjito

Afiahayati; Dwi AA Nugrahaningsih; Dyah A Puspitarani; Edwin W. Daniwijaya; Eggi Arguni; Endah Supriyati; Fadila D T Utami; Gita C Gabriela; Gunadi; Hendra Wibawa; Ika Trisnawati; Khanza A Vujira; Kristy Iskandar; Lanang Aditama; Laudria S Eryvinka; Ludhang P. Rizki; Mohamad S. Hakim; Nungki Anggorowati; Riat E Khair; Siswanto; Titik Nuryastuti; Tri Wibawa

Afiahayati; Dwi AA Nugrahaningsih; Dwi Indaryati; Dyah A Puspitarani; Edwin W. Daniwijaya; Eggi Arguni; Endah Supriyati; Fadila D T Utami; Gita C Gabriela; Gunadi; Hendra Wibawa; Ika Trisnawati; Indarto Sulistiyono; Khanza A Vujira; Kristy Iskandar; Lanang Aditama; Laudria S Eryvinka; Ludhang P. Rizki; Mohamad S. Hakim; Nungki Anggorowati; Pramesti G Dewi; Riat E Khair; Siswanto; Titik Nuryastuti; Tri Wibawa

EPI_ISL_10639785	Lab RS Ortopedi Prof. DR.R Soeharso Surakarta	Genetics Working Group (Pokja Genetik) Faculty of Medicine, Public Health and Nursing Universitas Gadjah Mada (FK-KMK UGM);

Disease Investigation Center Wates Ministry of Agriculture Indonesia; Department of Microbiology FK-KMK UGM; Laboratorium Diagnostik Yayasan Tahija World Mosquito Program (WMP) Yogyakarta Center for Tropical Medicine FK-KMK UGM; Integrated Research Center FK-KMK UGM; Department of Computer Science and Electronics FMIPA UGM; RSUP Dr. Sardjito

EPI_ISL_9713703	Lab RS Panti Rapih	Genetics Working Group (Pokja Genetik) Faculty of Medicine, Public Health and Nursing Universitas Gadjah Mada (FK-KMK UGM);

Disease Investigation Center Wates Ministry of Agriculture Indonesia; Department of Microbiology FK-KMK UGM; Laboratorium

Afiahayati; Dwi AA Nugrahaningsih; Dyah A Puspitarani; Edita M Devana; Edwin W. Daniwijaya; Eggi Arguni; Endah Supriyati; Esensi T Geometri; Fadila D T Utami; Gita C Gabriela; Gunadi; Hendra Wibawa; Ika Trisnawati; Khanza A Vujira; Kristy Iskandar; Ludhang P. Rizki; Mohamad S. Hakim; Nungki Anggorowati; Riat E Khair; Siswanto; Titik Nuryastuti; Tri Wibawa; Verrell Christopher

Afiahayati; Dwi AA Nugrahaningsih; Dyah A Puspitarani; Edwin W. Daniwijaya; Eggi Arguni; Endah Supriyati; Fadila D T Utami; Gita C Gabriela; Gunadi; Hendra Wibawa; Ika Trisnawati; Khanza A Vujira; Kristy Iskandar; Lanang Aditama; Laudria S Eryvinka; Ludhang P. Rizki; Mohamad S. Hakim; Nungki Anggorowati; Riat E Khair; Siswanto; Titik Nuryastuti; Tri Wibawa

EPI_ISL_7160967, EPI_ISL_7160969, EPI_ISL_7160974, EPI_ISL_7160979

Diagnostik Yayasan Tahija World Mosquito Program (WMP) Yogyakarta Center for Tropical Medicine FK-KMK UGM; Integrated Research Center FK-KMK UGM; Department of Computer Science and Electronics FMIPA UGM; RSUP Dr. Sardjito

Labkes Bantul	Genetics Working Group (Pokja Genetik) Faculty of Medicine, Public Health and Nursing Universitas Gadjah Mada (FK-KMK UGM); Disease Investigation Center Wates Ministry of Agriculture Indonesia; Department of Microbiology FK-KMK UGM; Laboratorium Diagnostik Yayasan Tahija World Mosquito Program (WMP) Yogyakarta Center for Tropical Medicine FK-KMK UGM; Integrated

Research Center FK-KMK UGM; Department of Computer Science and Electronics FMIPA UGM; RSUP Dr. Sardjito

Afiahayati; Dwi AA Nugrahaningsih; Dwi Indaryati; Dyah A Puspitarani; Edwin W. Daniwijaya; Eggi Arguni; Endah Supriyati; Gita C Gabriela; Gunadi; Hendra Wibawa; Indarto Sulistiyono; Khanza A Vujira; Khanza a Vujira; Kristy Iskandar; Ludhang P. Rizki; Marcellus; Mohamad S. Hakim; Nungki Anggorowati; Pramesti G Dewi; Siswanto; Titik Nuryastuti; Tri Wibawa

EPI_ISL_7154369, EPI_ISL_7160160, EPI_ISL_7160261, EPI_ISL_7160378, EPI_ISL_9487413, EPI_ISL_9713706, EPI_ISL_9713707

see above	Labkesda Bantul	Genetics Working Group (Pokja Genetik) Faculty of Medicine, Public Health and Nursing Universitas Gadjah Mada (FK-KMK UGM);

Disease Investigation Center Wates Ministry of Agriculture Indonesia; Department of Microbiology FK-KMK UGM; Laboratorium Diagnostik Yayasan Tahija World Mosquito Program (WMP) Yogyakarta Center for Tropical Medicine FK-KMK UGM; Integrated Research Center FK-KMK UGM; Department of Computer Science and Electronics FMIPA UGM; RSUP Dr. Sardjito

EPI_ISL_4969011	Labkesda Bantul DIY	Genetics Working Group (Pokja Genetik) Faculty of Medicine, Public Health and Nursing Universitas Gadjah Mada (FK-KMK UGM);

Disease Investigation Center Wates Ministry of Agriculture Indonesia; Department of Microbiology FK-KMK UGM; Laboratorium Diagnostik Yayasan Tahija World Mosquito Program (WMP) Yogyakarta Center for Tropical Medicine FK-KMK UGM; Integrated Research Center FK-KMK UGM; Department of Computer Science and Electronics FMIPA UGM; RSUP Dr. Sardjito

Afiahayati; Dwi AA Nugrahaningsih; Dwi Indaryati; Dyah A Purpitarani; Dyah A Puspitarani; Edwin W. Daniwijaya; Eggi Arguni; Endah Supriyati; Fadila D T Utami; Gita C Gabriela; Gunadi; Hendra Wibawa; Ika Trisnawati; Indarto Sulistiyono; Khanza A Vujira; Khanza a Vujira; Kristy Iskandar; Lanang Aditama; Lanang Aditama; Laudria S Eryvinka; Ludhang P. Rizki; Marcellus; Mohamad S. Hakim; Nungki Anggorowati; Pramesti G Dewi; Riat E Khair; Siswanto; Susan Simanjaya; Titik Nuryastuti; Tri Wibawa

Afiahayati; Alvina A Setiawan; Cita S Amalia; Dwi AA Nugrahaningsih; Dwi Indaryati; Edwin W. Daniwijaya; Eggi Arguni; Endah Supriyati; Gunadi; Hendra Wibawa; Indarto Sulistiyono; Kristy Iskandar; Ludhang P. Rizki; Marcellus; Mohamad S. Hakim; Nungki Anggorowati; Pramesti G Dewi; Siswanto; Titik Nuryastuti; Tri Wibawa

EPI_ISL_9487461, EPI_ISL_9487462, EPI_ISL_9487463

EPI_ISL_9702357, EPI_ISL_9755990, EPI_ISL_9756034

Labkesda Bantul/Puskesmas Imogiri I Bantul	Genetics Working Group (Pokja Genetik) Faculty of Medicine, Public Health and Nursing Universitas Gadjah Mada (FK-KMK UGM);

Disease Investigation Center Wates Ministry of Agriculture Indonesia; Department of Microbiology FK-KMK UGM; Laboratorium Diagnostik Yayasan Tahija World Mosquito Program (WMP) Yogyakarta Center for Tropical Medicine FK-KMK UGM; Integrated Research Center FK-KMK UGM; Department of Computer Science and Electronics FMIPA UGM; RSUP Dr. Sardjito

PUSKESMAS MLATI 1 SLEMAN	Genetics Working Group (Pokja Genetik) Faculty of Medicine, Public Health and Nursing Universitas Gadjah Mada (FK-KMK UGM); Disease Investigation Center Wates Ministry of Agriculture Indonesia; Department of Microbiology FK-KMK UGM; Laboratorium Diagnostik Yayasan Tahija World Mosquito Program (WMP) Yogyakarta Center for Tropical Medicine FK-KMK UGM; Integrated

Research Center FK-KMK UGM; Department of Computer Science and Electronics FMIPA UGM; RSUP Dr. Sardjito

Afiahayati; Dwi AA Nugrahaningsih; Dyah A Puspitarani; Edwin W. Daniwijaya; Eggi Arguni; Endah Supriyati; Fadila D T Utami; Gita C Gabriela; Gunadi; Hendra Wibawa; Ika Trisnawati; Khanza A Vujira; Kristy Iskandar; Lanang Aditama; Laudria S Eryvinka; Ludhang P. Rizki; Mohamad S. Hakim; Nungki Anggorowati; Riat E Khair; Siswanto; Titik Nuryastuti; Tri Wibawa

Afiahayati; Dwi AA Nugrahaningsih; Dyah A Puspitarani; Edwin W. Daniwijaya; Eggi Arguni; Endah Supriyati; Fadila D T Utami; Gita C Gabriela; Gunadi; Hendra Wibawa; Ika Trisnawati; Khanza A Vujira; Kristy Iskandar; Lanang Aditama; Laudria S Eryvinka; Ludhang P. Rizki; Mohamad S. Hakim; Nungki Anggorowati; Riat E Khair; Siswanto; Titik Nuryastuti; Tri Wibawa

EPI_ISL_9713705	PUSKESMAS Prambanan	Genetics Working Group (Pokja Genetik) Faculty of Medicine, Public Health and Nursing Universitas Gadjah Mada (FK-KMK UGM);

Disease Investigation Center Wates Ministry of Agriculture Indonesia; Department of Microbiology FK-KMK UGM; Laboratorium Diagnostik Yayasan Tahija World Mosquito Program (WMP) Yogyakarta Center for Tropical Medicine FK-KMK UGM; Integrated Research Center FK-KMK UGM; Department of Computer Science and Electronics FMIPA UGM; RSUP Dr. Sardjito

Afiahayati; Dwi AA Nugrahaningsih; Dyah A Puspitarani; Edwin W. Daniwijaya; Eggi Arguni; Endah Supriyati; Fadila D T Utami; Gita C Gabriela; Gunadi; Hendra Wibawa; Ika Trisnawati; Khanza A Vujira; Kristy Iskandar; Lanang Aditama; Laudria S Eryvinka; Ludhang P. Rizki; Mohamad S. Hakim; Nungki Anggorowati; Riat E Khair; Siswanto; Titik Nuryastuti; Tri Wibawa

EPI_ISL_9755818, EPI_ISL_9755829

Puskesmas Cangkringan	Genetics Working Group (Pokja Genetik) Faculty of Medicine, Public Health and Nursing Universitas Gadjah Mada (FK-KMK UGM); Disease Investigation Center Wates Ministry of Agriculture Indonesia; Department of Microbiology FK-KMK UGM; Laboratorium Diagnostik Yayasan Tahija World Mosquito Program (WMP) Yogyakarta Center for Tropical Medicine FK-KMK UGM; Integrated

Research Center FK-KMK UGM; Department of Computer Science and Electronics FMIPA UGM; RSUP Dr. Sardjito

Afiahayati; Dwi AA Nugrahaningsih; Dyah A Puspitarani; Edwin W. Daniwijaya; Eggi Arguni; Endah Supriyati; Fadila D T Utami; Gita C Gabriela; Gunadi; Hendra Wibawa; Ika Trisnawati; Khanza A Vujira; Kristy Iskandar; Lanang Aditama; Laudria S Eryvinka; Ludhang P. Rizki; Mohamad S. Hakim; Nungki Anggorowati; Riat E Khair; Siswanto; Titik Nuryastuti; Tri Wibawa

EPI_ISL_4969122, EPI_ISL_4969127, EPI_ISL_4969136, EPI_ISL_4969298, EPI_ISL_4969302, EPI_ISL_5010083, EPI_ISL_5010110, EPI_ISL_5010321

see above	Puskesmas Galur I	Genetics Working Group (Pokja Genetik) Faculty of Medicine, Public Health and Nursing Universitas Gadjah Mada (FK-KMK UGM);

Disease Investigation Center Wates Ministry of Agriculture Indonesia; Department of Microbiology FK-KMK UGM; Laboratorium Diagnostik Yayasan Tahija World Mosquito Program (WMP) Yogyakarta Center for Tropical Medicine FK-KMK UGM; Integrated Research Center FK-KMK UGM; Department of Computer Science and Electronics FMIPA UGM; RSUP Dr. Sardjito

Afiahayati; Alvina A Setiawan; Cita S Amalia; Dwi AA Nugrahaningsih; Dwi Indaryati; Dyah A Puspitarani; Edwin W. Daniwijaya; Eggi Arguni; Endah Supriyati; Gunadi; Hana F Hanifin; Hendra Wibawa; Indarto Sulistiyono; Irene Tania; Khanza a Vujira; Kristy Iskandar; Ludhang P. Rizki; Marcellus; Mohamad S. Hakim; Nungki Anggorowati; Pramesti G Dewi; Siswanto; Susan Simanjaya; Titik Nuryastuti; Tri Wibawa

EPI_ISL_9755988, EPI_ISL_9755989

Puskesmas Gondokusuman	Genetics Working Group (Pokja Genetik) Faculty of Medicine, Public Health and Nursing Universitas Gadjah Mada (FK-KMK UGM); Disease Investigation Center Wates Ministry of Agriculture Indonesia; Department of Microbiology FK-KMK UGM; Laboratorium Diagnostik Yayasan Tahija World Mosquito Program (WMP) Yogyakarta Center for Tropical Medicine FK-KMK UGM; Integrated

Research Center FK-KMK UGM; Department of Computer Science and Electronics FMIPA UGM; RSUP Dr. Sardjito

Afiahayati; Dwi AA Nugrahaningsih; Dyah A Puspitarani; Edwin W. Daniwijaya; Eggi Arguni; Endah Supriyati; Fadila D T Utami; Gita C Gabriela; Gunadi; Hendra Wibawa; Ika Trisnawati; Khanza A Vujira; Kristy Iskandar; Lanang Aditama; Laudria S Eryvinka; Ludhang P. Rizki; Mohamad S. Hakim; Nungki Anggorowati; Riat E Khair; Siswanto; Titik Nuryastuti; Tri Wibawa

EPI_ISL_4982801	Puskesmas Grimulyo I	Genetics Working Group (Pokja Genetik) Faculty of Medicine, Public Health and Nursing Universitas Gadjah Mada (FK-KMK UGM);

Disease Investigation Center Wates Ministry of Agriculture Indonesia; Department of Microbiology FK-KMK UGM; Laboratorium Diagnostik Yayasan Tahija World Mosquito Program (WMP) Yogyakarta Center for Tropical Medicine FK-KMK UGM; Integrated Research Center FK-KMK UGM; Department of Computer Science and Electronics FMIPA UGM; RSUP Dr. Sardjito

EPI_ISL_5010106	Puskesmas Kalibawang	Genetics Working Group (Pokja Genetik) Faculty of Medicine, Public Health and Nursing Universitas Gadjah Mada (FK-KMK UGM);

Disease Investigation Center Wates Ministry of Agriculture Indonesia; Department of Microbiology FK-KMK UGM; Laboratorium Diagnostik Yayasan Tahija World Mosquito Program (WMP) Yogyakarta Center for Tropical Medicine FK-KMK UGM; Integrated Research Center FK-KMK UGM; Department of Computer Science and Electronics FMIPA UGM; RSUP Dr. Sardjito

Afiahayati; Dwi AA Nugrahaningsih; Dwi Indaryati; Edwin W. Daniwijaya; Eggi Arguni; Endah Supriyati; Gunadi; Hana F Hanifin; Hendra Wibawa; Indarto Sulistiyono; Irene Tania; Kristy Iskandar; Ludhang P. Rizki; Marcellus; Mohamad S. Hakim; Nungki Anggorowati; Pramesti G Dewi; Siswanto; Titik Nuryastuti; Tri Wibawa

Afiahayati; Alvina A Setiawan; Cita S Amalia; Dwi AA Nugrahaningsih; Dwi Indaryati; Edwin W. Daniwijaya; Eggi Arguni; Endah Supriyati; Gunadi; Hendra Wibawa; Indarto Sulistiyono; Kristy Iskandar; Ludhang P. Rizki; Marcellus; Mohamad S. Hakim; Nungki Anggorowati; Pramesti G Dewi; Siswanto; Titik Nuryastuti; Tri Wibawa

EPI_ISL_4983057, EPI_ISL_5010024, EPI_ISL_5010104, EPI_ISL_5010355

Puskesmas Kokap II	Genetics Working Group (Pokja Genetik) Faculty of Medicine, Public Health and Nursing Universitas Gadjah Mada (FK-KMK UGM); Disease Investigation Center Wates Ministry of Agriculture Indonesia; Department of Microbiology FK-KMK UGM; Laboratorium Diagnostik Yayasan Tahija World Mosquito Program (WMP) Yogyakarta Center for Tropical Medicine FK-KMK UGM; Integrated

Research Center FK-KMK UGM; Department of Computer Science and Electronics FMIPA UGM; RSUP Dr. Sardjito

Afiahayati; Alvina A Setiawan; Cita S Amalia; Dwi AA Nugrahaningsih; Dwi Indaryati; Dyah A Puspitarani; Edwin W. Daniwijaya; Eggi Arguni; Endah Supriyati; Gunadi; Hana F Hanifin; Hendra Wibawa; Indarto Sulistiyono; Irene Tania; Kristy Iskandar; Ludhang P. Rizki; Marcellus; Mohamad S. Hakim; Nungki Anggorowati; Pramesti G Dewi; Siswanto; Susan Simanjaya; Titik Nuryastuti; Tri Wibawa

EPI_ISL_5010073	Puskesmas Lendah I	Genetics Working Group (Pokja Genetik) Faculty of Medicine, Public Health and Nursing Universitas Gadjah Mada (FK-KMK UGM);

Disease Investigation Center Wates Ministry of Agriculture Indonesia; Department of Microbiology FK-KMK UGM; Laboratorium Diagnostik Yayasan Tahija World Mosquito Program (WMP) Yogyakarta Center for Tropical Medicine FK-KMK UGM; Integrated Research Center FK-KMK UGM; Department of Computer Science and Electronics FMIPA UGM; RSUP Dr. Sardjito

EPI_ISL_5010492	Puskesmas PLERET	Genetics Working Group (Pokja Genetik) Faculty of Medicine, Public Health and Nursing Universitas Gadjah Mada (FK-KMK UGM);

Disease Investigation Center Wates Ministry of Agriculture Indonesia; Department of Microbiology FK-KMK UGM; Laboratorium Diagnostik Yayasan Tahija World Mosquito Program (WMP) Yogyakarta Center for Tropical Medicine FK-KMK UGM; Integrated Research Center FK-KMK UGM; Department of Computer Science and Electronics FMIPA UGM; RSUP Dr. Sardjito

EPI_ISL_4980593	Puskesmas Pengasih I	Genetics Working Group (Pokja Genetik) Faculty of Medicine, Public Health and Nursing Universitas Gadjah Mada (FK-KMK UGM);

Disease Investigation Center Wates Ministry of Agriculture Indonesia; Department of Microbiology FK-KMK UGM; Laboratorium Diagnostik Yayasan Tahija World Mosquito Program (WMP) Yogyakarta Center for Tropical Medicine FK-KMK UGM; Integrated Research Center FK-KMK UGM; Department of Computer Science and Electronics FMIPA UGM; RSUP Dr. Sardjito

Afiahayati; Alvina A Setiawan; Cita S Amalia; Dwi AA Nugrahaningsih; Dwi Indaryati; Edwin W. Daniwijaya; Eggi Arguni; Endah Supriyati; Gunadi; Hendra Wibawa; Indarto Sulistiyono; Kristy Iskandar; Ludhang P. Rizki; Marcellus; Mohamad S. Hakim; Nungki Anggorowati; Pramesti G Dewi; Siswanto; Titik Nuryastuti; Tri Wibawa

Afiahayati; Alvina A Setiawan; Dwi AA Nugrahaningsih; Dwi Indaryati; Edwin W. Daniwijaya; Eggi Arguni; Endah Supriyati; Gunadi; Hendra Wibawa; Indarto Sulistiyono; Khanza a Vujira; Kristy Iskandar; Ludhang P. Rizki; Marcellus; Mohamad S. Hakim; Nungki Anggorowati; Pramesti G Dewi; Siswanto; Titik Nuryastuti; Tri Wibawa

Afiahayati; Alvina A Setiawan; Dwi AA Nugrahaningsih; Dwi Indaryati; Edwin W. Daniwijaya; Eggi Arguni; Endah Supriyati; Gunadi; Hendra Wibawa; Indarto Sulistiyono; Khanza a Vujira; Kristy Iskandar; Ludhang P. Rizki; Marcellus; Mohamad S. Hakim; Nungki Anggorowati; Pramesti G Dewi; Siswanto; Titik Nuryastuti; Tri Wibawa

EPI_ISL_4980431, EPI_ISL_4983391

Puskesmas Pengasih II	Genetics Working Group (Pokja Genetik) Faculty of Medicine, Public Health and Nursing Universitas Gadjah Mada (FK-KMK UGM); Disease Investigation Center Wates Ministry of Agriculture Indonesia; Department of Microbiology FK-KMK UGM; Laboratorium Diagnostik Yayasan Tahija World Mosquito Program (WMP) Yogyakarta Center for Tropical Medicine FK-KMK UGM; Integrated

Research Center FK-KMK UGM; Department of Computer Science and Electronics FMIPA UGM; RSUP Dr. Sardjito

Afiahayati; Alvina A Setiawan; Cita S Amalia; Dwi AA Nugrahaningsih; Dwi Indaryati; Edwin W. Daniwijaya; Eggi Arguni; Endah Supriyati; Gunadi; Hendra Wibawa; Indarto Sulistiyono; Khanza a Vujira; Kristy Iskandar; Ludhang P. Rizki; Marcellus; Mohamad S. Hakim; Nungki Anggorowati; Pramesti G Dewi; Siswanto; Titik Nuryastuti; Tri Wibawa

EPI_ISL_5010416	Puskesmas Pleret	Genetics Working Group (Pokja Genetik) Faculty of Medicine, Public Health and Nursing Universitas Gadjah Mada (FK-KMK UGM);

Disease Investigation Center Wates Ministry of Agriculture Indonesia; Department of Microbiology FK-KMK UGM; Laboratorium Diagnostik Yayasan Tahija World Mosquito Program (WMP) Yogyakarta Center for Tropical Medicine FK-KMK UGM; Integrated Research Center FK-KMK UGM; Department of Computer Science and Electronics FMIPA UGM; RSUP Dr. Sardjito

EPI_ISL_5010063	Puskesmas Samigaluh I	Genetics Working Group (Pokja Genetik) Faculty of Medicine, Public Health and Nursing Universitas Gadjah Mada (FK-KMK UGM);

Disease Investigation Center Wates Ministry of Agriculture Indonesia; Department of Microbiology FK-KMK UGM; Laboratorium Diagnostik Yayasan Tahija World Mosquito Program (WMP) Yogyakarta Center for Tropical Medicine FK-KMK UGM; Integrated Research Center FK-KMK UGM; Department of Computer Science and Electronics FMIPA UGM; RSUP Dr. Sardjito

EPI_ISL_4969291	Puskesmas Temon 2	Genetics Working Group (Pokja Genetik) Faculty of Medicine, Public Health and Nursing Universitas Gadjah Mada (FK-KMK UGM);

Disease Investigation Center Wates Ministry of Agriculture Indonesia; Department of Microbiology FK-KMK UGM; Laboratorium Diagnostik Yayasan Tahija World Mosquito Program (WMP) Yogyakarta Center for Tropical Medicine FK-KMK UGM; Integrated Research Center FK-KMK UGM; Department of Computer Science and Electronics FMIPA UGM; RSUP Dr. Sardjito

EPI_ISL_7163388, EPI_ISL_7163917, EPI_ISL_7166591, EPI_ISL_7166628, EPI_ISL_7166791, EPI_ISL_7167384, EPI_ISL_7167528

see above	RS Akademik UGM	Genetics Working Group (Pokja Genetik) Faculty of Medicine, Public Health and Nursing Universitas Gadjah Mada (FK-KMK UGM);

Disease Investigation Center Wates Ministry of Agriculture Indonesia; Department of Microbiology FK-KMK UGM; Laboratorium Diagnostik Yayasan Tahija World Mosquito Program (WMP) Yogyakarta Center for Tropical Medicine FK-KMK UGM; Integrated Research Center FK-KMK UGM; Department of Computer Science and Electronics FMIPA UGM; RSUP Dr. Sardjito

Afiahayati; Dwi AA Nugrahaningsih; Dwi Indaryati; Dyah A Puspitarani; Edwin W. Daniwijaya; Eggi Arguni; Endah Supriyati; Gunadi; Hendra Wibawa; Indarto Sulistiyono; Kristy Iskandar; Ludhang P. Rizki; Marcellus; Mohamad S. Hakim; Nungki Anggorowati; Pramesti G Dewi; Siswanto; Susan Simanjaya; Titik Nuryastuti; Tri Wibawa

Afiahayati; Dwi AA Nugrahaningsih; Dwi Indaryati; Dyah A Puspitarani; Edwin W. Daniwijaya; Eggi Arguni; Endah Supriyati; Gunadi; Hendra Wibawa; Indarto Sulistiyono; Kristy Iskandar; Ludhang P. Rizki; Marcellus; Mohamad S. Hakim; Nungki Anggorowati; Pramesti G Dewi; Siswanto; Susan Simanjaya; Titik Nuryastuti; Tri Wibawa

Afiahayati; Alvina A Setiawan; Dwi AA Nugrahaningsih; Dwi Indaryati; Edwin W. Daniwijaya; Eggi Arguni; Endah Supriyati; Gunadi; Hendra Wibawa; Indarto Sulistiyono; Khanza a Vujira; Kristy Iskandar; Ludhang P. Rizki; Marcellus; Mohamad S. Hakim; Nungki Anggorowati; Pramesti G Dewi; Siswanto; Titik Nuryastuti; Tri Wibawa

Afiahayati; Dwi AA Nugrahaningsih; Dwi Indaryati; Dyah A Puspitarani; Edwin W. Daniwijaya; Eggi Arguni; Endah Supriyati; Gita C Gabriela; Gunadi; Hendra Wibawa; Indarto Sulistiyono; Khanza A Vujira; Kristy Iskandar; Ludhang P. Rizki; Marcellus; Mohamad S. Hakim; Nungki Anggorowati; Pramesti G Dewi; Siswanto; Susan Simanjaya; Titik Nuryastuti; Tri Wibawa

EPI_ISL_10657734, EPI_ISL_10658766

RS DR. OEN KANDANG SAPI SOLO	Genetics Working Group (Pokja Genetik) Faculty of Medicine, Public Health and Nursing Universitas Gadjah Mada (FK-KMK UGM); Disease Investigation Center Wates Ministry of Agriculture Indonesia; Department of Microbiology FK-KMK UGM; Laboratorium Diagnostik Yayasan Tahija World Mosquito Program (WMP) Yogyakarta Center for Tropical Medicine FK-KMK UGM; Integrated

Research Center FK-KMK UGM; Department of Computer Science and Electronics FMIPA UGM; RSUP Dr. Sardjito

Afiahayati; Dwi AA Nugrahaningsih; Dyah A Puspitarani; Edita M Devana; Edwin W. Daniwijaya; Eggi Arguni; Endah Supriyati; Esensi T Geometri; Fadila D T Utami; Gita C Gabriela; Gunadi; Hendra Wibawa; Ika Trisnawati; Khanza A Vujira; Kristy Iskandar; Ludhang P. Rizki; Mohamad S. Hakim; Nungki Anggorowati; Riat E Khair; Siswanto; Titik Nuryastuti; Tri Wibawa; Verrell Christopher

EPI_ISL_11800694	RS Dr. Oen Solo Baru	Genetics Working Group (Pokja Genetik) Faculty of Medicine, Public Health and Nursing Universitas Gadjah Mada (FK-KMK UGM);

Disease Investigation Center Wates Ministry of Agriculture Indonesia; BBTKLPP Yogyakarta; Department of Microbiology FK-KMK UGM; Laboratorium Diagnostik Yayasan Tahija World Mosquito Program (WMP) Yogyakarta Center for Tropical Medicine FK-KMK UGM; Integrated Research Center FK-KMK UGM; Department of Computer Science and Electronics FMIPA UGM; RSUP Dr. Sardjito

Afiahayati; Aning; Dwi AA Nugrahaningsih; Dyah A Puspitarani; Edita M Devana; Edwin W. Daniwijaya; Eggi Arguni; Endah Supriyati; Esensi T Geometri; Fadila D T Utami; Fatin; Gita C Gabriela; Gunadi; Hani; Havid; Hendra Wibawa; Ika Trisnawati; Irene; Khanza A Vujira; Kristy Iskandar; Lanang Aditama; Laudria S Eryvinka; Mohamad S. Hakim; Nathania C P Kinasih; Rahmi Nanda; Riat E Khair; Siswanto; Sri Fatmawati; Titik Nuryastuti; Tri Wibawa; Verrell Christopher

EPI_ISL_10639787, EPI_ISL_10657733, EPI_ISL_10658765

RS Dr. Oen Solo Baru	Genetics Working Group (Pokja Genetik) Faculty of Medicine, Public Health and Nursing Universitas Gadjah Mada (FK-KMK UGM); Disease Investigation Center Wates Ministry of Agriculture Indonesia; Department of Microbiology FK-KMK UGM; Laboratorium Diagnostik Yayasan Tahija World Mosquito Program (WMP) Yogyakarta Center for Tropical Medicine FK-KMK UGM; Integrated

Research Center FK-KMK UGM; Department of Computer Science and Electronics FMIPA UGM; RSUP Dr. Sardjito

Afiahayati; Dwi AA Nugrahaningsih; Dyah A Puspitarani; Edita M Devana; Edwin W. Daniwijaya; Eggi Arguni; Endah Supriyati; Esensi T Geometri; Fadila D T Utami; Gita C Gabriela; Gunadi; Hendra Wibawa; Ika Trisnawati; Khanza A Vujira; Kristy Iskandar; Ludhang P. Rizki; Mohamad S. Hakim; Nungki Anggorowati; Riat E Khair; Siswanto; Titik Nuryastuti; Tri Wibawa; Verrell Christopher

EPI_ISL_4968087,	RS JIH	Genetics Working Group (Pokja Genetik) Faculty of Medicine, Public Health and Nursing Universitas Gadjah Mada (FK-KMK UGM); Afiahayati; Alvina A Setiawan; Cita S Amalia; Dwi AA Nugrahaningsih; Dwi Indaryati; Dyah A Puspitarani; Edwin W. Daniwijaya; Eggi Arguni; Endah Supriyati; Gunadi; Hana F Hanifin;

| EPI_ISL_4969010, EPI_ISL_4969082,  EPI_ISL_4969086 |  | Disease Investigation Center Wates Ministry of Agriculture Indonesia; Department of Microbiology FK-KMK UGM; Laboratorium Diagnostik Yayasan Tahija World Mosquito Program (WMP) Yogyakarta Center for Tropical Medicine FK-KMK UGM; Integrated  Research Center FK-KMK UGM; Department of Computer Science and Electronics FMIPA UGM; RSUP Dr. Sardjito | Hendra Wibawa; Indarto Sulistiyono; Irene Tania; Khanza a Vujira; Kristy Iskandar; Ludhang P. Rizki; Marcellus; Mohamad S. Hakim; Nungki Anggorowati; Pramesti G Dewi; Siswanto; Susan Simanjaya; Titik Nuryastuti; Tri Wibawa |
| --- | --- | --- | --- |
| EPI_ISL_10658764 | RS KASIH IBU | Genetics Working Group (Pokja Genetik) Faculty of Medicine, Public Health and Nursing Universitas Gadjah Mada (FK-KMK UGM); Disease Investigation Center Wates Ministry of Agriculture Indonesia; Department of Microbiology FK-KMK UGM; Laboratorium Diagnostik Yayasan Tahija World Mosquito Program (WMP) Yogyakarta Center for Tropical Medicine FK-KMK UGM; Integrated Research Center FK-KMK UGM; Department of Computer Science and Electronics FMIPA UGM; RSUP Dr. Sardjito | Afiahayati; Dwi AA Nugrahaningsih; Dyah A Puspitarani; Edita M Devana; Edwin W. Daniwijaya; Eggi Arguni; Endah Supriyati; Esensi T Geometri; Fadila D T Utami; Gita C Gabriela; Gunadi; Hendra Wibawa; Ika Trisnawati; Khanza A Vujira; Kristy Iskandar; Ludhang P. Rizki; Mohamad S. Hakim; Nungki Anggorowati; Riat E Khair; Siswanto; Titik Nuryastuti; Tri Wibawa; Verrell Christopher |
| EPI_ISL_10640044 | RS KASIH IBU SURAKARTA | Genetics Working Group (Pokja Genetik) Faculty of Medicine, Public Health and Nursing Universitas Gadjah Mada (FK-KMK UGM); Disease Investigation Center Wates Ministry of Agriculture Indonesia; Department of Microbiology FK-KMK UGM; Laboratorium Diagnostik Yayasan Tahija World Mosquito Program (WMP) Yogyakarta Center for Tropical Medicine FK-KMK UGM; Integrated Research Center FK-KMK UGM; Department of Computer Science and Electronics FMIPA UGM; RSUP Dr. Sardjito | Afiahayati; Dwi AA Nugrahaningsih; Dyah A Puspitarani; Edita M Devana; Edwin W. Daniwijaya; Eggi Arguni; Endah Supriyati; Esensi T Geometri; Fadila D T Utami; Gita C Gabriela; Gunadi; Hendra Wibawa; Ika Trisnawati; Khanza A Vujira; Kristy Iskandar; Ludhang P. Rizki; Mohamad S. Hakim; Nungki Anggorowati; Riat E Khair; Siswanto; Titik Nuryastuti; Tri Wibawa; Verrell Christopher |
| EPI_ISL_9755959 | RS PKU Gamping | Genetics Working Group (Pokja Genetik) Faculty of Medicine, Public Health and Nursing Universitas Gadjah Mada (FK-KMK UGM); Disease Investigation Center Wates Ministry of Agriculture Indonesia; Department of Microbiology FK-KMK UGM; Laboratorium Diagnostik Yayasan Tahija World Mosquito Program (WMP) Yogyakarta Center for Tropical Medicine FK-KMK UGM; Integrated Research Center FK-KMK UGM; Department of Computer Science and Electronics FMIPA UGM; RSUP Dr. Sardjito | Afiahayati; Dwi AA Nugrahaningsih; Dyah A Puspitarani; Edwin W. Daniwijaya; Eggi Arguni; Endah Supriyati; Fadila D T Utami; Gita C Gabriela; Gunadi; Hendra Wibawa; Ika Trisnawati; Khanza A Vujira; Kristy Iskandar; Lanang Aditama; Laudria S Eryvinka; Ludhang P. Rizki; Mohamad S. Hakim; Nungki Anggorowati; Riat E Khair; Siswanto; Titik Nuryastuti; Tri Wibawa |
| EPI_ISL_9713702 | RS PKU Kota Yogyakarta | Genetics Working Group (Pokja Genetik) Faculty of Medicine, Public Health and Nursing Universitas Gadjah Mada (FK-KMK UGM); Disease Investigation Center Wates Ministry of Agriculture Indonesia; Department of Microbiology FK-KMK UGM; Laboratorium Diagnostik Yayasan Tahija World Mosquito Program (WMP) Yogyakarta Center for Tropical Medicine FK-KMK UGM; Integrated Research Center FK-KMK UGM; Department of Computer Science and Electronics FMIPA UGM; RSUP Dr. Sardjito | Afiahayati; Dwi AA Nugrahaningsih; Dyah A Puspitarani; Edwin W. Daniwijaya; Eggi Arguni; Endah Supriyati; Fadila D T Utami; Gita C Gabriela; Gunadi; Hendra Wibawa; Ika Trisnawati; Khanza A Vujira; Kristy Iskandar; Lanang Aditama; Laudria S Eryvinka; Ludhang P. Rizki; Mohamad S. Hakim; Nungki Anggorowati; Riat E Khair; Siswanto; Titik Nuryastuti; Tri Wibawa |
| EPI_ISL_9713704 | RS PKU Muh. Gamping | Genetics Working Group (Pokja Genetik) Faculty of Medicine, Public Health and Nursing Universitas Gadjah Mada (FK-KMK UGM); Disease Investigation Center Wates Ministry of Agriculture Indonesia; Department of Microbiology FK-KMK UGM; Laboratorium Diagnostik Yayasan Tahija World Mosquito Program (WMP) Yogyakarta Center for Tropical Medicine FK-KMK UGM; Integrated Research Center FK-KMK UGM; Department of Computer Science and Electronics FMIPA UGM; RSUP Dr. Sardjito | Afiahayati; Dwi AA Nugrahaningsih; Dyah A Puspitarani; Edwin W. Daniwijaya; Eggi Arguni; Endah Supriyati; Fadila D T Utami; Gita C Gabriela; Gunadi; Hendra Wibawa; Ika Trisnawati; Khanza A Vujira; Kristy Iskandar; Lanang Aditama; Laudria S Eryvinka; Ludhang P. Rizki; Mohamad S. Hakim; Nungki Anggorowati; Riat E Khair; Siswanto; Titik Nuryastuti; Tri Wibawa |
| EPI_ISL_9487456 | RS PKU Muhammadiyah Bantul | Genetics Working Group (Pokja Genetik) Faculty of Medicine, Public Health and Nursing Universitas Gadjah Mada (FK-KMK UGM); Disease Investigation Center Wates Ministry of Agriculture Indonesia; Department of Microbiology FK-KMK UGM; Laboratorium Diagnostik Yayasan Tahija World Mosquito Program (WMP) Yogyakarta Center for Tropical Medicine FK-KMK UGM; Integrated Research Center FK-KMK UGM; Department of Computer Science and Electronics FMIPA UGM; RSUP Dr. Sardjito | Afiahayati; Dwi AA Nugrahaningsih; Dyah A Puspitarani; Edwin W. Daniwijaya; Eggi Arguni; Endah Supriyati; Fadila D T Utami; Gita C Gabriela; Gunadi; Hendra Wibawa; Ika Trisnawati; Khanza A Vujira; Kristy Iskandar; Lanang Aditama; Laudria S Eryvinka; Ludhang P. Rizki; Mohamad S. Hakim; Nungki Anggorowati; Riat E Khair; Siswanto; Titik Nuryastuti; Tri Wibawa |
| EPI_ISL_9433335, EPI_ISL_9434455 | RS Panti Rapih | Genetics Working Group (Pokja Genetik) Faculty of Medicine, Public Health and Nursing Universitas Gadjah Mada (FK-KMK UGM); Disease Investigation Center Wates Ministry of Agriculture Indonesia; Department of Microbiology FK-KMK UGM; Laboratorium Diagnostik Yayasan Tahija World Mosquito Program (WMP) Yogyakarta Center for Tropical Medicine FK-KMK UGM; Integrated Research Center FK-KMK UGM; Department of Computer Science and Electronics FMIPA UGM; RSUP Dr. Sardjito | Afiahayati; Dwi AA Nugrahaningsih; Dwi Indaryati; Dyah A Puspitarani; Edwin W. Daniwijaya; Eggi Arguni; Endah Supriyati; Gunadi; Hendra Wibawa; Indarto Sulistiyono; Khanza A Vujira; Kristy Iskandar; Ludhang P. Rizki; Mohamad S. Hakim; Nungki Anggorowati; Pramesti G Dewi; Siswanto; Titik Nuryastuti; Tri Wibawa |
| EPI_ISL_5010527 | RS Rajawali Citra | Genetics Working Group (Pokja Genetik) Faculty of Medicine, Public Health and Nursing Universitas Gadjah Mada (FK-KMK UGM); Disease Investigation Center Wates Ministry of Agriculture Indonesia; Department of Microbiology FK-KMK UGM; Laboratorium Diagnostik Yayasan Tahija World Mosquito Program (WMP) Yogyakarta Center for Tropical Medicine FK-KMK UGM; Integrated Research Center FK-KMK UGM; Department of Computer Science and Electronics FMIPA UGM; RSUP Dr. Sardjito | Afiahayati; Alvina A Setiawan; Cita S Amalia; Dwi AA Nugrahaningsih; Dwi Indaryati; Edwin W. Daniwijaya; Eggi Arguni; Endah Supriyati; Gunadi; Hendra Wibawa; Indarto Sulistiyono; Kristy Iskandar; Ludhang P. Rizki; Marcellus; Mohamad S. Hakim; Nungki Anggorowati; Pramesti G Dewi; Siswanto; Titik Nuryastuti; Tri Wibawa |
| EPI_ISL_10969463, EPI_ISL_10969464 | RS. Pertamina Cilacap | Genetics Working Group (Pokja Genetik) Faculty of Medicine, Public Health and Nursing Universitas Gadjah Mada (FK-KMK UGM); Balai Besar Teknik Kesehatan Lingkungan Dan Pengendalian Penyakit (BBTKLPP) Yogyakarta; Department of Microbiology FK-KMK UGM; Laboratorium Diagnostik Yayasan Tahija World Mosquito Program (WMP) Yogyakarta Center for Tropical Medicine FK-KMK UGM; Integrated Research Center FK-KMK UGM; Department of Computer Science and Electronics FMIPA UGM; RSUP Dr. Sardjito | Afiahayati; Aning; Dwi AA Nugrahaningsih; Dyah A Puspitarani; Edita M Devana; Edwin W. Daniwijaya; Eggi Arguni; Endah Supriyati; Esensi T Geometri; Fadila D T Utami; Fatin; Gita C Gabriela; Gunadi; Hani; Havid; Hendra Wibawa; Ika Trisnawati; Irene; Khanza A Vujira; Kristy Iskandar; Lanang Aditama; Laudria S Eryvinka; Mohamad S. Hakim; Nathania C P Kinasih; Rahmi Nanda; Riat E Khair; Siswanto; Sri Fatmawati; Titik Nuryastuti; Tri Wibawa; Verrell Christopher |
| EPI_ISL_10969465, EPI_ISL_10969466 | RSA UGM | Genetics Working Group (Pokja Genetik) Faculty of Medicine, Public Health and Nursing Universitas Gadjah Mada (FK-KMK UGM); Balai Besar Teknik Kesehatan Lingkungan Dan Pengendalian Penyakit (BBTKLPP) Yogyakarta; Department of Microbiology FK-KMK UGM; Laboratorium Diagnostik Yayasan Tahija World Mosquito Program (WMP) Yogyakarta Center for Tropical Medicine FK-KMK UGM; Integrated Research Center FK-KMK UGM; Department of Computer Science and Electronics FMIPA UGM; RSUP Dr. Sardjito | Afiahayati; Aning; Dwi AA Nugrahaningsih; Dyah A Puspitarani; Edita M Devana; Edwin W. Daniwijaya; Eggi Arguni; Endah Supriyati; Esensi T Geometri; Fadila D T Utami; Fatin; Gita C Gabriela; Gunadi; Hani; Havid; Hendra Wibawa; Ika Trisnawati; Irene; Khanza A Vujira; Kristy Iskandar; Lanang Aditama; Laudria S Eryvinka; Mohamad S. Hakim; Nathania C P Kinasih; Rahmi Nanda; Riat E Khair; Siswanto; Sri Fatmawati; Titik Nuryastuti; Tri Wibawa; Verrell Christopher |

EPI_ISL_7162211, EPI_ISL_7162387, EPI_ISL_7162539, EPI_ISL_7162961, EPI_ISL_7163289, EPI_ISL_7163308, EPI_ISL_7163385, EPI_ISL_9434592, EPI_ISL_9434818, EPI_ISL_9487464, EPI_ISL_12241066

see above	RSA UGM	Genetics Working Group (Pokja Genetik) Faculty of Medicine, Public Health and Nursing Universitas Gadjah Mada (FK-KMK UGM);

Disease Investigation Center Wates Ministry of Agriculture Indonesia; Department of Microbiology FK-KMK UGM; Laboratorium Diagnostik Yayasan Tahija World Mosquito Program (WMP) Yogyakarta Center for Tropical Medicine FK-KMK UGM; Integrated Research Center FK-KMK UGM; Department of Computer Science and Electronics FMIPA UGM; RSUP Dr. Sardjito

EPI_ISL_4969084	RSI NU Demak	Genetics Working Group (Pokja Genetik) Faculty of Medicine, Public Health and Nursing Universitas Gadjah Mada (FK-KMK UGM);

Disease Investigation Center Wates Ministry of Agriculture Indonesia; Department of Microbiology FK-KMK UGM; Laboratorium Diagnostik Yayasan Tahija World Mosquito Program (WMP) Yogyakarta Center for Tropical Medicine FK-KMK UGM; Integrated Research Center FK-KMK UGM; Department of Computer Science and Electronics FMIPA UGM; RSUP Dr. Sardjito

EPI_ISL_7166965	RSKIA Sadewa	Genetics Working Group (Pokja Genetik) Faculty of Medicine, Public Health and Nursing Universitas Gadjah Mada (FK-KMK UGM);

Disease Investigation Center Wates Ministry of Agriculture Indonesia; Department of Microbiology FK-KMK UGM; Laboratorium Diagnostik Yayasan Tahija World Mosquito Program (WMP) Yogyakarta Center for Tropical Medicine FK-KMK UGM; Integrated Research Center FK-KMK UGM; Department of Computer Science and Electronics FMIPA UGM; RSUP Dr. Sardjito

EPI_ISL_9487465	RSPAU dr. S.Hardjolukito	Genetics Working Group (Pokja Genetik) Faculty of Medicine, Public Health and Nursing Universitas Gadjah Mada (FK-KMK UGM);

Disease Investigation Center Wates Ministry of Agriculture Indonesia; Department of Microbiology FK-KMK UGM; Laboratorium Diagnostik Yayasan Tahija World Mosquito Program (WMP) Yogyakarta Center for Tropical Medicine FK-KMK UGM; Integrated Research Center FK-KMK UGM; Department of Computer Science and Electronics FMIPA UGM; RSUP Dr. Sardjito

Afiahayati; Dwi AA Nugrahaningsih; Dwi Indaryati; Dyah A Puspitarani; Edwin W. Daniwijaya; Eggi Arguni; Endah Supriyati; Fadila D T Utami; Gita C Gabriela; Gunadi; Hendra Wibawa; Ika Trisnawati; Indarto Sulistiyono; Khanza A Vujira; Kristy Iskandar; Lanang Aditama; Laudria S Eryvinka; Ludhang P. Rizki; Marcellus; Mohamad S. Hakim; Nungki Anggorowati; Pramesti G Dewi; Riat E Khair; Siswanto; Susan Simanjaya; Titik Nuryastuti; Tri Wibawa

Afiahayati; Alvina A Setiawan; Dwi AA Nugrahaningsih; Dwi Indaryati; Edwin W. Daniwijaya; Eggi Arguni; Endah Supriyati; Gunadi; Hendra Wibawa; Indarto Sulistiyono; Khanza a Vujira; Kristy Iskandar; Ludhang P. Rizki; Marcellus; Mohamad S. Hakim; Nungki Anggorowati; Pramesti G Dewi; Siswanto; Titik Nuryastuti; Tri Wibawa

Afiahayati; Dwi AA Nugrahaningsih; Dwi Indaryati; Dyah A Puspitarani; Edwin W. Daniwijaya; Eggi Arguni; Endah Supriyati; Gita C Gabriela; Gunadi; Hendra Wibawa; Indarto Sulistiyono; Khanza A Vujira; Kristy Iskandar; Ludhang P. Rizki; Marcellus; Mohamad S. Hakim; Nungki Anggorowati; Pramesti G Dewi; Siswanto; Titik Nuryastuti; Tri Wibawa

Afiahayati; Dwi AA Nugrahaningsih; Dyah A Puspitarani; Edwin W. Daniwijaya; Eggi Arguni; Endah Supriyati; Fadila D T Utami; Gita C Gabriela; Gunadi; Hendra Wibawa; Ika Trisnawati; Khanza A Vujira; Kristy Iskandar; Lanang Aditama; Laudria S Eryvinka; Ludhang P. Rizki; Mohamad S. Hakim; Nungki Anggorowati; Riat E Khair; Siswanto; Titik Nuryastuti; Tri Wibawa

EPI_ISL_9702414, EPI_ISL_9702417, EPI_ISL_9713700, EPI_ISL_9713701

RSPAU dr.S.Hardjolukito	Genetics Working Group (Pokja Genetik) Faculty of Medicine, Public Health and Nursing Universitas Gadjah Mada (FK-KMK UGM); Disease Investigation Center Wates Ministry of Agriculture Indonesia; Department of Microbiology FK-KMK UGM; Laboratorium Diagnostik Yayasan Tahija World Mosquito Program (WMP) Yogyakarta Center for Tropical Medicine FK-KMK UGM; Integrated

Research Center FK-KMK UGM; Department of Computer Science and Electronics FMIPA UGM; RSUP Dr. Sardjito

Afiahayati; Dwi AA Nugrahaningsih; Dyah A Puspitarani; Edwin W. Daniwijaya; Eggi Arguni; Endah Supriyati; Fadila D T Utami; Gita C Gabriela; Gunadi; Hendra Wibawa; Ika Trisnawati; Khanza A Vujira; Kristy Iskandar; Lanang Aditama; Laudria S Eryvinka; Ludhang P. Rizki; Mohamad S. Hakim; Nungki Anggorowati; Riat E Khair; Siswanto; Titik Nuryastuti; Tri Wibawa

EPI_ISL_7160962	RSST Klaten	Genetics Working Group (Pokja Genetik) Faculty of Medicine, Public Health and Nursing Universitas Gadjah Mada (FK-KMK UGM);

Disease Investigation Center Wates Ministry of Agriculture Indonesia; Department of Microbiology FK-KMK UGM; Laboratorium Diagnostik Yayasan Tahija World Mosquito Program (WMP) Yogyakarta Center for Tropical Medicine FK-KMK UGM; Integrated Research Center FK-KMK UGM; Department of Computer Science and Electronics FMIPA UGM; RSUP Dr. Sardjito

Afiahayati; Dwi AA Nugrahaningsih; Dwi Indaryati; Dyah A Puspitarani; Edwin W. Daniwijaya; Eggi Arguni; Endah Supriyati; Gita C Gabriela; Gunadi; Hendra Wibawa; Indarto Sulistiyono; Khanza a Vujira; Kristy Iskandar; Ludhang P. Rizki; Marcellus; Mohamad S. Hakim; Nungki Anggorowati; Pramesti G Dewi; Siswanto; Titik Nuryastuti; Tri Wibawa

EPI_ISL_10969451, EPI_ISL_10969452, EPI_ISL_10969453, EPI_ISL_10969454, EPI_ISL_10969455

EPI_ISL_10969460, EPI_ISL_10969461, EPI_ISL_10969462

EPI_ISL_10640043, EPI_ISL_12240910

RSU AISYIYAH	Genetics Working Group (Pokja Genetik) Faculty of Medicine, Public Health and Nursing Universitas Gadjah Mada (FK-KMK UGM); Balai Besar Teknik Kesehatan Lingkungan Dan Pengendalian Penyakit (BBTKLPP) Yogyakarta; Department of Microbiology FK-KMK

UGM; Laboratorium Diagnostik Yayasan Tahija World Mosquito Program (WMP) Yogyakarta Center for Tropical Medicine FK-KMK UGM; Integrated Research Center FK-KMK UGM; Department of Computer Science and Electronics FMIPA UGM; RSUP Dr. Sardjito

RSUD Wonogiri	Genetics Working Group (Pokja Genetik) Faculty of Medicine, Public Health and Nursing Universitas Gadjah Mada (FK-KMK UGM); Balai Besar Teknik Kesehatan Lingkungan Dan Pengendalian Penyakit (BBTKLPP) Yogyakarta; Department of Microbiology FK-KMK

UGM; Laboratorium Diagnostik Yayasan Tahija World Mosquito Program (WMP) Yogyakarta Center for Tropical Medicine FK-KMK UGM; Integrated Research Center FK-KMK UGM; Department of Computer Science and Electronics FMIPA UGM; RSUP Dr. Sardjito

RSUD Ibu Fatmawati Soekarno	Genetics Working Group (Pokja Genetik) Faculty of Medicine, Public Health and Nursing Universitas Gadjah Mada (FK-KMK UGM); Disease Investigation Center Wates Ministry of Agriculture Indonesia; Department of Microbiology FK-KMK UGM; Laboratorium Diagnostik Yayasan Tahija World Mosquito Program (WMP) Yogyakarta Center for Tropical Medicine FK-KMK UGM; Integrated

Research Center FK-KMK UGM; Department of Computer Science and Electronics FMIPA UGM; RSUP Dr. Sardjito

Afiahayati; Aning; Dwi AA Nugrahaningsih; Dyah A Puspitarani; Edita M Devana; Edwin W. Daniwijaya; Eggi Arguni; Endah Supriyati; Esensi T Geometri; Fadila D T Utami; Fatin; Gita C Gabriela; Gunadi; Hani; Havid; Hendra Wibawa; Ika Trisnawati; Irene; Khanza A Vujira; Kristy Iskandar; Lanang Aditama; Laudria S Eryvinka; Mohamad S. Hakim; Nathania C P Kinasih; Rahmi Nanda; Riat E Khair; Siswanto; Sri Fatmawati; Titik Nuryastuti; Tri Wibawa; Verrell Christopher

Afiahayati; Aning; Dwi AA Nugrahaningsih; Dyah A Puspitarani; Edita M Devana; Edwin W. Daniwijaya; Eggi Arguni; Endah Supriyati; Esensi T Geometri; Fadila D T Utami; Fatin; Gita C Gabriela; Gunadi; Hani; Havid; Hendra Wibawa; Ika Trisnawati; Irene; Khanza A Vujira; Kristy Iskandar; Lanang Aditama; Laudria S Eryvinka; Mohamad S. Hakim; Nathania C P Kinasih; Rahmi Nanda; Riat E Khair; Siswanto; Sri Fatmawati; Titik Nuryastuti; Tri Wibawa; Verrell Christopher

Afiahayati; Dwi AA Nugrahaningsih; Dyah A Puspitarani; Edita M Devana; Edwin W. Daniwijaya; Eggi Arguni; Endah Supriyati; Esensi T Geometri; Fadila D T Utami; Gita C Gabriela; Gunadi; Hendra Wibawa; Ika Trisnawati; Khanza A Vujira; Kristy Iskandar; Ludhang P. Rizki; Mohamad S. Hakim; Nungki Anggorowati; Riat E Khair; Siswanto; Titik Nuryastuti; Tri Wibawa; Verrell Christopher

EPI_ISL_7160960, EPI_ISL_7160961, EPI_ISL_7160982, EPI_ISL_7160983, EPI_ISL_7161078, EPI_ISL_7161365, EPI_ISL_7161601, EPI_ISL_7161977

see above	RSUD K.M.R.T Wongsonegoro	Genetics Working Group (Pokja Genetik) Faculty of Medicine, Public Health and Nursing Universitas Gadjah Mada (FK-KMK UGM);

Disease Investigation Center Wates Ministry of Agriculture Indonesia; Department of Microbiology FK-KMK UGM; Laboratorium Diagnostik Yayasan Tahija World Mosquito Program (WMP) Yogyakarta Center for Tropical Medicine FK-KMK UGM; Integrated Research Center FK-KMK UGM; Department of Computer Science and Electronics FMIPA UGM; RSUP Dr. Sardjito

. Marcellus; Afiahayati; Bambang Sigit Riyanto; Dwi AA Nugrahaningsih; Dwi Indaryati; Dyah A Puspitarani; Edwin W. Daniwijaya; Eggi Arguni; Eko Budiono; Endah Supriyati; Gita C Gabriela; Gita C Gabriela; Gunadi; Hendra Wibawa; Heni Retnowulan; Ika Trisnawati; Indarto Sulistiyono; Ira Puspitawati; Khanza A Vujira; Khanza a Vujira; Kristy Iskandar; Ludhang P. Rizki; Marcellus; Mohamad S. Hakim; Munawar Gani; Nungki Anggorowati; Nur Imma Fatimah Harahap; Nur Rahmi Ananda; Osman Sianipar; Pramesti G Dewi; Riat El Khair; Satria Maulana; Siswanto; Sumardi; Susan Simanjaya; Titik Nuryastuti; Tri Wibawa; Umi Solekhah Intansari; Yunika Puspadewi; Elizabeth Henny Herningtiyas

EPI_ISL_10639788, EPI_ISL_10639789, EPI_ISL_10657581, EPI_ISL_10658763, EPI_ISL_10658769

EPI_ISL_4969091, EPI_ISL_4969096, EPI_ISL_4969115, EPI_ISL_4969119

EPI_ISL_10969449, EPI_ISL_10969450, EPI_ISL_10969456, EPI_ISL_10969457,

RSUD KRT. SETJONEGORO	Genetics Working Group (Pokja Genetik) Faculty of Medicine, Public Health and Nursing Universitas Gadjah Mada (FK-KMK UGM); Disease Investigation Center Wates Ministry of Agriculture Indonesia; Department of Microbiology FK-KMK UGM; Laboratorium Diagnostik Yayasan Tahija World Mosquito Program (WMP) Yogyakarta Center for Tropical Medicine FK-KMK UGM; Integrated

Research Center FK-KMK UGM; Department of Computer Science and Electronics FMIPA UGM; RSUP Dr. Sardjito

RSUD KRT. Setjonegoro Wonosobo	Genetics Working Group (Pokja Genetik) Faculty of Medicine, Public Health and Nursing Universitas Gadjah Mada (FK-KMK UGM); Disease Investigation Center Wates Ministry of Agriculture Indonesia; Department of Microbiology FK-KMK UGM; Laboratorium Diagnostik Yayasan Tahija World Mosquito Program (WMP) Yogyakarta Center for Tropical Medicine FK-KMK UGM; Integrated

Research Center FK-KMK UGM; Department of Computer Science and Electronics FMIPA UGM; RSUP Dr. Sardjito

RSUD MUNTILAN	Genetics Working Group (Pokja Genetik) Faculty of Medicine, Public Health and Nursing Universitas Gadjah Mada (FK-KMK UGM); Balai Besar Teknik Kesehatan Lingkungan Dan Pengendalian Penyakit (BBTKLPP) Yogyakarta; Department of Microbiology FK-KMK

UGM; Laboratorium Diagnostik Yayasan Tahija World Mosquito Program (WMP) Yogyakarta Center for Tropical Medicine FK-KMK UGM; Integrated Research Center FK-KMK UGM; Department of Computer Science and Electronics FMIPA UGM; RSUP Dr. Sardjito

Afiahayati; Dwi AA Nugrahaningsih; Dyah A Puspitarani; Edita M Devana; Edwin W. Daniwijaya; Eggi Arguni; Endah Supriyati; Esensi T Geometri; Fadila D T Utami; Gita C Gabriela; Gunadi; Hendra Wibawa; Ika Trisnawati; Khanza A Vujira; Kristy Iskandar; Ludhang P. Rizki; Mohamad S. Hakim; Nungki Anggorowati; Riat E Khair; Siswanto; Titik Nuryastuti; Tri Wibawa; Verrell Christopher

. Marcellus; Afiahayati; Alvina A Setiawan; Bambang Sigit Riyanto; Dwi AA Nugrahaningsih; Dwi Indaryati; Dyah A Puspitarani; Edwin W. Daniwijaya; Eggi Arguni; Eko Budiono; Endah Supriyati; Gunadi; Hana F Hanifin; Hendra Wibawa; Heni Retnowulan; Ika Trisnawati; Indarto Sulistiyono; Ira Puspitawati; Irene Tania; Khanza a Vujira; Kristy Iskandar; Ludhang P. Rizki; Marcellus; Mohamad S. Hakim; Munawar Gani; Nungki Anggorowati; Nur Imma Fatimah Harahap; Nur Rahmi Ananda; Osman Sianipar; Pramesti G Dewi; Riat El Khair; Satria Maulana; Siswanto; Sumardi; Susan Simanjaya; Titik Nuryastuti; Tri Wibawa; Umi Solekhah Intansari; Yunika Puspadewi; Elizabeth Henny Herningtiyas

Afiahayati; Aning; Dwi AA Nugrahaningsih; Dyah A Puspitarani; Edita M Devana; Edwin W. Daniwijaya; Eggi Arguni; Endah Supriyati; Esensi T Geometri; Fadila D T Utami; Fatin; Gita C Gabriela; Gunadi; Hani; Havid; Hendra Wibawa; Ika Trisnawati; Irene; Khanza A Vujira; Kristy Iskandar; Lanang Aditama; Laudria S Eryvinka; Mohamad S. Hakim; Nathania C P Kinasih; Rahmi Nanda; Riat E Khair; Siswanto; Sri Fatmawati; Titik Nuryastuti; Tri Wibawa; Verrell Christopher

EPI_ISL_10969458, EPI_ISL_10969459

EPI_ISL_4978455, EPI_ISL_4980931, EPI_ISL_4981198, EPI_ISL_4981563, EPI_ISL_5010016, EPI_ISL_5010036, EPI_ISL_5010038, EPI_ISL_5010046, EPI_ISL_5010054, EPI_ISL_5010412

see above	RSUD Nyi Ageng Serang	Genetics Working Group (Pokja Genetik) Faculty of Medicine, Public Health and Nursing Universitas Gadjah Mada (FK-KMK UGM);

Disease Investigation Center Wates Ministry of Agriculture Indonesia; Department of Microbiology FK-KMK UGM; Laboratorium Diagnostik Yayasan Tahija World Mosquito Program (WMP) Yogyakarta Center for Tropical Medicine FK-KMK UGM; Integrated Research Center FK-KMK UGM; Department of Computer Science and Electronics FMIPA UGM; RSUP Dr. Sardjito

. Marcellus; Afiahayati; Alvina A Setiawan; Bambang Sigit Riyanto; Cita S Amalia; Dwi AA Nugrahaningsih; Dwi Indaryati; Dyah A Puspitarani; Edwin W. Daniwijaya; Eggi Arguni; Eko Budiono; Endah Supriyati; Gunadi; Hana F Hanifin; Hendra Wibawa; Heni Retnowulan; I Putu Aditio A; Ika Trisnawati; Indarto Sulistiyono; Ira Puspitawati; Irene Tania; Khanza a Vujira; Kristy Iskandar; Ludhang P. Rizki; Marcellus; Mohamad S. Hakim; Munawar Gani; Nungki Anggorowati; Nur Imma Fatimah Harahap; Nur Rahmi Ananda; Osman Sianipar; Pramesti G Dewi; Riat El Khair; Satria Maulana; Siswanto; Sumardi; Susan Simanjaya; Titik Nuryastuti; Tri Wibawa; Umi Solekhah Intansari; Yunika Puspadewi; Elizabeth Henny Herningtiyas

EPI_ISL_4969177, EPI_ISL_4969179

EPI_ISL_7153225, EPI_ISL_7153586

RSUD Sunan Kalijaga Demak	Genetics Working Group (Pokja Genetik) Faculty of Medicine, Public Health and Nursing Universitas Gadjah Mada (FK-KMK UGM); Disease Investigation Center Wates Ministry of Agriculture Indonesia; Department of Microbiology FK-KMK UGM; Laboratorium Diagnostik Yayasan Tahija World Mosquito Program (WMP) Yogyakarta Center for Tropical Medicine FK-KMK UGM; Integrated

Research Center FK-KMK UGM; Department of Computer Science and Electronics FMIPA UGM; RSUP Dr. Sardjito

RSUD dr. Loekmono Hadi	Genetics Working Group (Pokja Genetik) Faculty of Medicine, Public Health and Nursing Universitas Gadjah Mada (FK-KMK UGM); Disease Investigation Center Wates Ministry of Agriculture Indonesia; Department of Microbiology FK-KMK UGM; Laboratorium Diagnostik Yayasan Tahija World Mosquito Program (WMP) Yogyakarta Center for Tropical Medicine FK-KMK UGM; Integrated

Research Center FK-KMK UGM; Department of Computer Science and Electronics FMIPA UGM; RSUP Dr. Sardjito

Afiahayati; Alvina A Setiawan; Dwi AA Nugrahaningsih; Dwi Indaryati; Dyah A Puspitarani; Edwin W. Daniwijaya; Eggi Arguni; Endah Supriyati; Gunadi; Hendra Wibawa; Indarto Sulistiyono; Khanza a Vujira; Kristy Iskandar; Ludhang P. Rizki; Marcellus; Mohamad S. Hakim; Nungki Anggorowati; Pramesti G Dewi; Siswanto; Susan Simanjaya; Titik Nuryastuti; Tri Wibawa

Afiahayati; Dwi AA Nugrahaningsih; Dwi Indaryati; Dyah A Puspitarani; Edwin W. Daniwijaya; Eggi Arguni; Endah Supriyati; Gita C Gabriela; Gunadi; Hendra Wibawa; Indarto Sulistiyono; Khanza A Vujira; Kristy Iskandar; Ludhang P. Rizki; Marcellus; Mohamad S. Hakim; Nungki Anggorowati; Pramesti G Dewi; Siswanto; Susan Simanjaya; Titik Nuryastuti; Tri Wibawa

EPI_ISL_4969112	RSUP Dr Sardjito	Genetics Working Group (Pokja Genetik) Faculty of Medicine, Public Health and Nursing Universitas Gadjah Mada (FK-KMK UGM);

Disease Investigation Center Wates Ministry of Agriculture Indonesia; Department of Microbiology FK-KMK UGM; Laboratorium Diagnostik Yayasan Tahija World Mosquito Program (WMP) Yogyakarta Center for Tropical Medicine FK-KMK UGM; Integrated Research Center FK-KMK UGM; Department of Computer Science and Electronics FMIPA UGM; RSUP Dr. Sardjito

Afiahayati; Dwi AA Nugrahaningsih; Dwi Indaryati; Edwin W. Daniwijaya; Eggi Arguni; Endah Supriyati; Gunadi; Hana F Hanifin; Hendra Wibawa; Indarto Sulistiyono; Irene Tania; Kristy Iskandar; Ludhang P. Rizki; Marcellus; Mohamad S. Hakim; Nungki Anggorowati; Pramesti G Dewi; Siswanto; Titik Nuryastuti; Tri Wibawa

EPI_ISL_10626964, EPI_ISL_10626972, EPI_ISL_10626973, EPI_ISL_10626974, EPI_ISL_10626975, EPI_ISL_10626976, EPI_ISL_10626977, EPI_ISL_10626978, EPI_ISL_10626979, EPI_ISL_10626980, EPI_ISL_10626981, EPI_ISL_10627013, EPI_ISL_10627014

see above	RSUP Dr. Sardjito	Genetics Working Group (Pokja Genetik) Faculty of Medicine, Public Health and Nursing Universitas Gadjah Mada (FK-KMK UGM);

Disease Investigation Center Wates Ministry of Agriculture Indonesia; Department of Microbiology FK-KMK UGM; Laboratorium Diagnostik Yayasan Tahija World Mosquito Program (WMP) Yogyakarta Center for Tropical Medicine FK-KMK UGM; Integrated Research Center FK-KMK UGM; Department of Computer Science and Electronics FMIPA UGM; RSUP Dr. Sardjito

Afiahayati; Dwi AA Nugrahaningsih; Dyah A Puspitarani; Edita M Devana; Edwin W. Daniwijaya; Eggi Arguni; Endah Supriyati; Esensi T Geometri; Fadila D T Utami; Gita C Gabriela; Gunadi; Hendra Wibawa; Ika Trisnawati; Khanza A Vujira; Kristy Iskandar; Ludhang P. Rizki; Mohamad S. Hakim; Nungki Anggorowati; Riat E Khair; Siswanto; Titik Nuryastuti; Tri Wibawa; Verrell Christopher

EPI_ISL_10969467, EPI_ISL_10969468, EPI_ISL_10969469, EPI_ISL_10969470, EPI_ISL_10969471, EPI_ISL_10969472, EPI_ISL_10969473, EPI_ISL_10969474, EPI_ISL_10969475, EPI_ISL_10969476, EPI_ISL_10969477, EPI_ISL_10969481, EPI_ISL_10969503, EPI_ISL_10969509, EPI_ISL_10969521, EPI_ISL_10969612, EPI_ISL_10969613, EPI_ISL_10969845, EPI_ISL_10970235, EPI_ISL_10970479, EPI_ISL_10977347, EPI_ISL_10977423, EPI_ISL_10977817, EPI_ISL_10978119, EPI_ISL_10978329, EPI_ISL_10978528, EPI_ISL_10979009, EPI_ISL_10979174

see above	RSUP dr. Sardjito	Genetics Working Group (Pokja Genetik) Faculty of Medicine, Public Health and Nursing Universitas Gadjah Mada (FK-KMK UGM);

Balai Besar Teknik Kesehatan Lingkungan Dan Pengendalian Penyakit (BBTKLPP) Yogyakarta; Department of Microbiology FK-KMK UGM; Laboratorium Diagnostik Yayasan Tahija World Mosquito Program (WMP) Yogyakarta Center for Tropical Medicine FK-KMK UGM; Integrated Research Center FK-KMK UGM; Department of Computer Science and Electronics FMIPA UGM; RSUP Dr. Sardjito

Afiahayati; Aning; Dwi AA Nugrahaningsih; Dyah A Puspitarani; Edita M Devana; Edwin W. Daniwijaya; Eggi Arguni; Endah Supriyati; Esensi T Geometri; Fadila D T Utami; Fatin; Gita C Gabriela; Gunadi; Hani; Havid; Hendra Wibawa; Ika Trisnawati; Irene; Khanza A Vujira; Kristy Iskandar; Lanang; Lanang Aditama; Laudria S Eryvinka; Ludhang P. Rizki; Mohamad S. Hakim; Nathania; Nathania C P Kinasih; Nungki Anggorowati; Rahmi Nanda; Riat E Khair; Siswanto; Sri Fatmawati; Stella; Titik Nuryastuti; Tri Wibawa; Verrell Christopher

EPI_ISL_9570723, EPI_ISL_9570724, EPI_ISL_9570726, EPI_ISL_9570728

RSUP dr. Sardjito	Genetics Working Group (Pokja Genetik) Faculty of Medicine, Public Health and Nursing Universitas Gadjah Mada (FK-KMK UGM); Disease Investigation Center Wates Ministry of Agriculture Indonesia; Department of Microbiology FK-KMK UGM; Laboratorium Diagnostik Yayasan Tahija World Mosquito Program (WMP) Yogyakarta Center for Tropical Medicine FK-KMK UGM; Integrated

Research Center FK-KMK UGM; Department of Computer Science and Electronics FMIPA UGM; RSUP Dr. Sardjito

Afiahayati; Dwi AA Nugrahaningsih; Dyah A Puspitarani; Edwin W. Daniwijaya; Eggi Arguni; Endah Supriyati; Fadila D T Utami; Gita C Gabriela; Gunadi; Hendra Wibawa; Ika Trisnawati; Khanza A Vujira; Kristy Iskandar; Lanang Aditama; Laudria S Eryvinka; Ludhang P. Rizki; Mohamad S. Hakim; Nungki Anggorowati; Riat E Khair; Siswanto; Titik Nuryastuti; Tri Wibawa

EPI_ISL_4968082	Rumah Sakit Bhayangkara Polda DIY	Genetics Working Group (Pokja Genetik) Faculty of Medicine, Public Health and Nursing Universitas Gadjah Mada (FK-KMK UGM);

Disease Investigation Center Wates Ministry of Agriculture Indonesia; Department of Microbiology FK-KMK UGM; Laboratorium Diagnostik Yayasan Tahija World Mosquito Program (WMP) Yogyakarta Center for Tropical Medicine FK-KMK UGM; Integrated Research Center FK-KMK UGM; Department of Computer Science and Electronics FMIPA UGM; RSUP Dr. Sardjito

Afiahayati; Dwi AA Nugrahaningsih; Dwi Indaryati; Edwin W. Daniwijaya; Eggi Arguni; Endah Supriyati; Gunadi; Hana F Hanifin; Hendra Wibawa; Indarto Sulistiyono; Irene Tania; Kristy Iskandar; Ludhang P. Rizki; Marcellus; Mohamad S. Hakim; Nungki Anggorowati; Pramesti G Dewi; Siswanto; Titik Nuryastuti; Tri Wibawa

EPI_ISL_10657577, EPI_ISL_10657727, EPI_ISL_10657728, EPI_ISL_10657729

Rumah Sakit JIH SOLO - The Ultimate Value Healthcare Genetics Working Group (Pokja Genetik) Faculty of Medicine, Public Health and Nursing Universitas Gadjah Mada (FK-KMK UGM);

Disease Investigation Center Wates Ministry of Agriculture Indonesia; Department of Microbiology FK-KMK UGM; Laboratorium Diagnostik Yayasan Tahija World Mosquito Program (WMP) Yogyakarta Center for Tropical Medicine FK-KMK UGM; Integrated Research Center FK-KMK UGM; Department of Computer Science and Electronics FMIPA UGM; RSUP Dr. Sardjito

Afiahayati; Dwi AA Nugrahaningsih; Dyah A Puspitarani; Edita M Devana; Edwin W. Daniwijaya; Eggi Arguni; Endah Supriyati; Esensi T Geometri; Fadila D T Utami; Gita C Gabriela; Gunadi; Hendra Wibawa; Ika Trisnawati; Khanza A Vujira; Kristy Iskandar; Ludhang P. Rizki; Mohamad S. Hakim; Nungki Anggorowati; Riat E Khair; Siswanto; Titik Nuryastuti; Tri Wibawa; Verrell Christopher

EPI_ISL_10658792	Rumah Sakit JIH Solo	Genetics Working Group (Pokja Genetik) Faculty of Medicine, Public Health and Nursing Universitas Gadjah Mada (FK-KMK UGM);

Disease Investigation Center Wates Ministry of Agriculture Indonesia; Department of Microbiology FK-KMK UGM; Laboratorium Diagnostik Yayasan Tahija World Mosquito Program (WMP) Yogyakarta Center for Tropical Medicine FK-KMK UGM; Integrated Research Center FK-KMK UGM; Department of Computer Science and Electronics FMIPA UGM; RSUP Dr. Sardjito

Afiahayati; Dwi AA Nugrahaningsih; Dyah A Puspitarani; Edita M Devana; Edwin W. Daniwijaya; Eggi Arguni; Endah Supriyati; Esensi T Geometri; Fadila D T Utami; Gita C Gabriela; Gunadi; Hendra Wibawa; Ika Trisnawati; Khanza A Vujira; Kristy Iskandar; Ludhang P. Rizki; Mohamad S. Hakim; Nungki Anggorowati; Riat E Khair; Siswanto; Titik Nuryastuti; Tri Wibawa; Verrell Christopher

EPI_ISL_10657579, EPI_ISL_10657731, EPI_ISL_10658770

EPI_ISL_9487326, EPI_ISL_9487383, EPI_ISL_9487411

Rumah Sakit Kasih Ibu Surakarta	Genetics Working Group (Pokja Genetik) Faculty of Medicine, Public Health and Nursing Universitas Gadjah Mada (FK-KMK UGM); Disease Investigation Center Wates Ministry of Agriculture Indonesia; Department of Microbiology FK-KMK UGM; Laboratorium Diagnostik Yayasan Tahija World Mosquito Program (WMP) Yogyakarta Center for Tropical Medicine FK-KMK UGM; Integrated

Research Center FK-KMK UGM; Department of Computer Science and Electronics FMIPA UGM; RSUP Dr. Sardjito

UPTD Labkes Kab. Kebumen	Genetics Working Group (Pokja Genetik) Faculty of Medicine, Public Health and Nursing Universitas Gadjah Mada (FK-KMK UGM); Disease Investigation Center Wates Ministry of Agriculture Indonesia; Department of Microbiology FK-KMK UGM; Laboratorium Diagnostik Yayasan Tahija World Mosquito Program (WMP) Yogyakarta Center for Tropical Medicine FK-KMK UGM; Integrated

Research Center FK-KMK UGM; Department of Computer Science and Electronics FMIPA UGM; RSUP Dr. Sardjito

Afiahayati; Dwi AA Nugrahaningsih; Dyah A Puspitarani; Edita M Devana; Edwin W. Daniwijaya; Eggi Arguni; Endah Supriyati; Esensi T Geometri; Fadila D T Utami; Gita C Gabriela; Gunadi; Hendra Wibawa; Ika Trisnawati; Khanza A Vujira; Kristy Iskandar; Ludhang P. Rizki; Mohamad S. Hakim; Nungki Anggorowati; Riat E Khair; Siswanto; Titik Nuryastuti; Tri Wibawa; Verrell Christopher

Afiahayati; Dwi AA Nugrahaningsih; Dyah A Puspitarani; Edwin W. Daniwijaya; Eggi Arguni; Endah Supriyati; Fadila D T Utami; Gita C Gabriela; Gunadi; Hendra Wibawa; Ika Trisnawati; Khanza A Vujira; Kristy Iskandar; Lanang Aditama; Laudria S Eryvinka; Ludhang P. Rizki; Mohamad S. Hakim; Nungki Anggorowati; Riat E Khair; Siswanto; Titik Nuryastuti; Tri Wibawa

### We gratefully acknowledge the following Authors from the Originating laboratories responsible for obtaining the specimens, as well as the Submitting laboratories where the genome data were generated and shared via GISAID, on which this research is based.

All Submitters of data may be contacted directly via [www.gisaid.org](http://www.gisaid.org/) Authors are sorted alphabetically.

#### Accession ID Originating Laboratory Submitting Laboratory Authors

EPI_ISL_2943235	BBTKLPP Yogyakarta	Genetics Working Group (Pokja Genetik) Faculty of Medicine, Public Health and Nursing Universitas Gadjah Mada (FK-KMK UGM); Disease

Investigation Center Wates Ministry of Agriculture Indonesia; Department of Microbiology FK-KMK UGM; Laboratorium Diagnostik Yayasan Tahija World Mosquito Program (WMP) Yogyakarta Center for Tropical Medicine FK-KMK UGM; Integrated Research Center FK-KMK UGM; Department of Computer Science and Electronics FMIPA UGM; RSUP Dr. Sardjito

Afiahayati; Dwi AA Nugrahaningsih; Dwi Indaryati; Edwin W. Daniwijaya; Eggi Arguni; Endah Supriyati; Gunadi; Hana F Hanifin; Hendra Wibawa; Indarto Sulistiyono; Irene Tania; Kristy Iskandar; Ludhang P. Rizki; Marcellus; Mohamad S. Hakim; Nungki Anggorowati; Pramesti G Dewi; Siswanto; Titik Nuryastuti; Tri Wibawa

EPI_ISL_2964704, EPI_ISL_2964706

EPI_ISL_2964697, EPI_ISL_2964700

DINKES KAB. GROBOGAN	Genetics Working Group (Pokja Genetik) Faculty of Medicine, Public Health and Nursing Universitas Gadjah Mada (FK-KMK UGM); Disease Investigation Center Wates Ministry of Agriculture Indonesia; Department of Microbiology FK-KMK UGM; Laboratorium Diagnostik Yayasan Tahija World Mosquito Program (WMP) Yogyakarta Center for Tropical Medicine FK-KMK UGM; Integrated Research Center FK-KMK UGM; Department of

Computer Science and Electronics FMIPA UGM; RSUP Dr. Sardjito

DINKES KAB. JEPARA	Genetics Working Group (Pokja Genetik) Faculty of Medicine, Public Health and Nursing Universitas Gadjah Mada (FK-KMK UGM); Disease Investigation Center Wates Ministry of Agriculture Indonesia; Department of Microbiology FK-KMK UGM; Laboratorium Diagnostik Yayasan Tahija World Mosquito Program (WMP) Yogyakarta Center for Tropical Medicine FK-KMK UGM; Integrated Research Center FK-KMK UGM; Department of

Computer Science and Electronics FMIPA UGM; RSUP Dr. Sardjito

Afiahayati; Dwi AA Nugrahaningsih; Dwi Indaryati; Dyah A Puspitarani; Edwin W. Daniwijaya; Eggi Arguni; Endah Supriyati; Gunadi; Hana F Hanifin; Hendra Wibawa; Indarto Sulistiyono; Irene Tania; Kristy Iskandar; Ludhang P. Rizki; Marcellus; Mohamad S. Hakim; Nungki Anggorowati; Pramesti G Dewi; Siswanto; Susan Simanjaya; Titik Nuryastuti; Tri Wibawa

Afiahayati; Alvina A Setiawan; Cita S Amalia; Dwi AA Nugrahaningsih; Dwi Indaryati; Edwin W. Daniwijaya; Eggi Arguni; Endah Supriyati; Gunadi; Hendra Wibawa; Indarto Sulistiyono; Khanza a Vujira; Kristy Iskandar; Ludhang P. Rizki; Marcellus; Mohamad S. Hakim; Nungki Anggorowati; Pramesti G Dewi; Siswanto; Titik Nuryastuti; Tri Wibawa

EPI_ISL_2954684	DKK Gunungkidul/ Puskesmas Girisubo	Genetics Working Group (Pokja Genetik) Faculty of Medicine, Public Health and Nursing Universitas Gadjah Mada (FK-KMK UGM); Disease

Investigation Center Wates Ministry of Agriculture Indonesia; Department of Microbiology FK-KMK UGM; Laboratorium Diagnostik Yayasan Tahija World Mosquito Program (WMP) Yogyakarta Center for Tropical Medicine FK-KMK UGM; Integrated Research Center FK-KMK UGM; Department of Computer Science and Electronics FMIPA UGM; RSUP Dr. Sardjito

Afiahayati; Alvina A Setiawan; Cita S Amalia; Dwi AA Nugrahaningsih; Dwi Indaryati; Edwin W. Daniwijaya; Eggi Arguni; Endah Supriyati; Gunadi; Hendra Wibawa; Indarto Sulistiyono; Kristy Iskandar; Ludhang P. Rizki; Marcellus; Mohamad S. Hakim; Nungki Anggorowati; Pramesti G Dewi; Siswanto; Titik Nuryastuti; Tri Wibawa

EPI_ISL_2955335, EPI_ISL_2955586, EPI_ISL_2955597, EPI_ISL_2955598

DKK Gunungkidul/ Puskesmas Karangmojo I	Genetics Working Group (Pokja Genetik) Faculty of Medicine, Public Health and Nursing Universitas Gadjah Mada (FK-KMK UGM); Disease Investigation Center Wates Ministry of Agriculture Indonesia; Department of Microbiology FK-KMK UGM; Laboratorium Diagnostik Yayasan Tahija World Mosquito Program (WMP) Yogyakarta Center for Tropical Medicine FK-KMK UGM; Integrated Research Center FK-KMK UGM; Department of

Computer Science and Electronics FMIPA UGM; RSUP Dr. Sardjito

Afiahayati; Alvina A Setiawan; Dwi AA Nugrahaningsih; Dwi Indaryati; Dyah A Puspitarani; Edwin W. Daniwijaya; Eggi Arguni; Endah Supriyati; Gunadi; Hana F Hanifin; Hendra Wibawa; Indarto Sulistiyono; Irene Tania; Khanza a Vujira; Kristy Iskandar; Ludhang P. Rizki; Marcellus; Mohamad S. Hakim; Nungki Anggorowati; Pramesti G Dewi; Siswanto; Susan Simanjaya; Titik Nuryastuti; Tri Wibawa

EPI_ISL_2933110	DKK Sleman/Puskesmas Depok 1	Genetics Working Group (Pokja Genetik) Faculty of Medicine, Public Health and Nursing Universitas Gadjah Mada (FK-KMK UGM); Disease

Investigation Center Wates Ministry of Agriculture Indonesia; Department of Microbiology FK-KMK UGM; Laboratorium Diagnostik Yayasan Tahija World Mosquito Program (WMP) Yogyakarta Center for Tropical Medicine FK-KMK UGM; Integrated Research Center FK-KMK UGM; Department of Computer Science and

Afiahayati; Alvina A Setiawan; Dwi AA Nugrahaningsih; Dwi Indaryati; Edwin W. Daniwijaya; Eggi Arguni; Endah Supriyati; Gunadi; Hendra Wibawa; Indarto Sulistiyono; Khanza a Vujira; Kristy Iskandar; Ludhang P. Rizki; Marcellus; Mohamad S. Hakim; Nungki Anggorowati; Pramesti G Dewi; Siswanto; Titik Nuryastuti; Tri Wibawa

EPI_ISL_2932613, EPI_ISL_2933109, EPI_ISL_2943237, EPI_ISL_2955609

EPI_ISL_2955627, EPI_ISL_2955628

DKK Sleman/Puskesmas Depok 1	Genetics Working Group (Pokja Genetik) Faculty of Medicine, Public Health and Nursing Universitas Gadjah Mada (FK-KMK UGM); Disease Investigation Center Wates Ministry of Agriculture Indonesia; Department of Microbiology FK-KMK UGM; Laboratorium Diagnostik Yayasan Tahija World Mosquito Program (WMP) Yogyakarta Center for Tropical Medicine FK-KMK UGM; Integrated Research Center FK-KMK UGM; Department of

Computer Science and Electronics FMIPA UGM; RSUP Dr. Sardjito

DKK Sleman/Puskesmas Ngemplak 2	Genetics Working Group (Pokja Genetik) Faculty of Medicine, Public Health and Nursing Universitas Gadjah Mada (FK-KMK UGM); Disease Investigation Center Wates Ministry of Agriculture Indonesia; Department of Microbiology FK-KMK UGM; Laboratorium Diagnostik Yayasan Tahija World Mosquito Program (WMP) Yogyakarta Center for Tropical Medicine FK-KMK UGM; Integrated Research Center FK-KMK UGM; Department of

Computer Science and Electronics FMIPA UGM; RSUP Dr. Sardjito

Afiahayati; Alvina A Setiawan; Cita S Amalia; Dwi AA Nugrahaningsih; Dwi Indaryati; Dyah A Puspitarani; Edwin W. Daniwijaya; Eggi Arguni; Endah Supriyati; Gunadi; Hana F Hanifin; Hendra Wibawa; Indarto Sulistiyono; Irene Tania; Kristy Iskandar; Ludhang P. Rizki; Marcellus; Mohamad S. Hakim; Nungki Anggorowati; Pramesti G Dewi; Siswanto; Susan Simanjaya; Titik Nuryastuti; Tri Wibawa

Afiahayati; Dwi AA Nugrahaningsih; Dwi Indaryati; Dyah A Puspitarani; Edwin W. Daniwijaya; Eggi Arguni; Endah Supriyati; Gunadi; Hana F Hanifin; Hendra Wibawa; Indarto Sulistiyono; Irene Tania; Kristy Iskandar; Ludhang P. Rizki; Marcellus; Mohamad S. Hakim; Nungki Anggorowati; Pramesti G Dewi; Siswanto; Susan Simanjaya; Titik Nuryastuti; Tri Wibawa

EPI_ISL_2932610	KOTA Yogya DKK Gondokusuman II	Genetics Working Group (Pokja Genetik) Faculty of Medicine, Public Health and Nursing Universitas Gadjah Mada (FK-KMK UGM); Disease

Investigation Center Wates Ministry of Agriculture Indonesia; Department of Microbiology FK-KMK UGM; Laboratorium Diagnostik Yayasan Tahija World Mosquito Program (WMP) Yogyakarta Center for

EPI_ISL_2964681	Puskesmas Colomadu 1 DKK Karanganyar	Genetics Working Group (Pokja Genetik) Faculty of Medicine, Public Health and Nursing Universitas Gadjah Mada (FK-KMK UGM); Disease

Investigation Center Wates Ministry of Agriculture Indonesia; Department of Microbiology FK-KMK UGM; Laboratorium Diagnostik Yayasan Tahija World Mosquito Program (WMP) Yogyakarta Center for Tropical Medicine FK-KMK UGM; Integrated Research Center FK-KMK UGM; Department of Computer Science and Electronics FMIPA UGM; RSUP Dr. Sardjito

Afiahayati; Alvina A Setiawan; Dwi AA Nugrahaningsih; Dwi Indaryati; Edwin W. Daniwijaya; Eggi Arguni; Endah Supriyati; Gunadi; Hendra Wibawa; Indarto Sulistiyono; Khanza a Vujira; Kristy Iskandar; Ludhang P. Rizki; Marcellus; Mohamad S. Hakim; Nungki Anggorowati; Pramesti G Dewi; Siswanto; Titik Nuryastuti; Tri Wibawa

Afiahayati; Alvina A Setiawan; Dwi AA Nugrahaningsih; Dwi Indaryati; Edwin W. Daniwijaya; Eggi Arguni; Endah Supriyati; Gunadi; Hendra Wibawa; Indarto Sulistiyono; Khanza a Vujira; Kristy Iskandar; Ludhang P. Rizki; Marcellus; Mohamad S. Hakim; Nungki Anggorowati; Pramesti G Dewi; Siswanto; Titik Nuryastuti; Tri Wibawa

EPI_ISL_2964917, EPI_ISL_2964918, EPI_ISL_2964919, EPI_ISL_2964920, EPI_ISL_2964921

EPI_ISL_2964707, EPI_ISL_2964872

RS Dr. OEN KANDANG SAPI SOLO	Genetics Working Group (Pokja Genetik) Faculty of Medicine, Public Health and Nursing Universitas Gadjah Mada (FK-KMK UGM); Disease Investigation Center Wates Ministry of Agriculture Indonesia; Department of Microbiology FK-KMK UGM; Laboratorium Diagnostik Yayasan Tahija World Mosquito Program (WMP) Yogyakarta Center for Tropical Medicine FK-KMK UGM; Integrated Research Center FK-KMK UGM; Department of

Computer Science and Electronics FMIPA UGM; RSUP Dr. Sardjito

RS Dr. Oen Kandang Sapi Solo	Genetics Working Group (Pokja Genetik) Faculty of Medicine, Public Health and Nursing Universitas Gadjah Mada (FK-KMK UGM); Disease Investigation Center Wates Ministry of Agriculture Indonesia; Department of Microbiology FK-KMK UGM; Laboratorium Diagnostik Yayasan Tahija World Mosquito Program (WMP) Yogyakarta Center for Tropical Medicine FK-KMK UGM; Integrated Research Center FK-KMK UGM; Department of

Computer Science and Electronics FMIPA UGM; RSUP Dr. Sardjito

Afiahayati; Alvina A Setiawan; Cita S Amalia; Dwi AA Nugrahaningsih; Dwi Indaryati; Dyah A Puspitarani; Edwin W. Daniwijaya; Eggi Arguni; Endah Supriyati; Gunadi; Hana F Hanifin; Hendra Wibawa; Indarto Sulistiyono; Irene Tania; Khanza a Vujira; Kristy Iskandar; Ludhang P. Rizki; Marcellus; Mohamad S. Hakim; Nungki Anggorowati; Pramesti G Dewi; Siswanto; Susan Simanjaya; Titik Nuryastuti; Tri Wibawa

Afiahayati; Alvina A Setiawan; Cita S Amalia; Dwi AA Nugrahaningsih; Dwi Indaryati; Dyah A Puspitarani; Edwin W. Daniwijaya; Eggi Arguni; Endah Supriyati; Gunadi; Hendra Wibawa; Indarto Sulistiyono; Kristy Iskandar; Ludhang P. Rizki; Marcellus; Mohamad S. Hakim; Nungki Anggorowati; Pramesti G Dewi; Siswanto; Susan Simanjaya; Titik Nuryastuti; Tri Wibawa

EPI_ISL_2955608	RSA UGM	Genetics Working Group (Pokja Genetik) Faculty of Medicine, Public Health and Nursing Universitas Gadjah Mada (FK-KMK UGM); Disease

Investigation Center Wates Ministry of Agriculture Indonesia; Department of Microbiology FK-KMK UGM; Laboratorium Diagnostik Yayasan Tahija World Mosquito Program (WMP) Yogyakarta Center for Tropical Medicine FK-KMK UGM; Integrated Research Center FK-KMK UGM; Department of Computer Science and Electronics FMIPA UGM; RSUP Dr. Sardjito

EPI_ISL_2964684, EPI_ISL_2964688, EPI_ISL_2964692, EPI_ISL_2964941, EPI_ISL_2964942, EPI_ISL_2964943, EPI_ISL_2964944, EPI_ISL_2964956, EPI_ISL_2964957

see above	RSUD DR. MOEWARDI	Genetics Working Group (Pokja Genetik) Faculty of Medicine, Public Health and Nursing Universitas Gadjah Mada (FK-KMK UGM); Disease

Investigation Center Wates Ministry of Agriculture Indonesia; Department of Microbiology FK-KMK UGM; Laboratorium Diagnostik Yayasan Tahija World Mosquito Program (WMP) Yogyakarta Center for Tropical Medicine FK-KMK UGM; Integrated Research Center FK-KMK UGM; Department of Computer Science and Electronics FMIPA UGM; RSUP Dr. Sardjito

EPI_ISL_2964677	RSUD DR. MOEWARDI Surakarta	Genetics Working Group (Pokja Genetik) Faculty of Medicine, Public Health and Nursing Universitas Gadjah Mada (FK-KMK UGM); Disease

Investigation Center Wates Ministry of Agriculture Indonesia; Department of Microbiology FK-KMK UGM; Laboratorium Diagnostik Yayasan Tahija World Mosquito Program (WMP) Yogyakarta Center for Tropical Medicine FK-KMK UGM; Integrated Research Center FK-KMK UGM; Department of Computer Science and Electronics FMIPA UGM; RSUP Dr. Sardjito

Afiahayati; Dwi AA Nugrahaningsih; Dwi Indaryati; Dyah A Puspitarani; Edwin W. Daniwijaya; Eggi Arguni; Endah Supriyati; Gunadi; Hendra Wibawa; Indarto Sulistiyono; Kristy Iskandar; Ludhang P. Rizki; Marcellus; Mohamad S. Hakim; Nungki Anggorowati; Pramesti G Dewi; Siswanto; Susan Simanjaya; Titik Nuryastuti; Tri Wibawa

Afiahayati; Alvina A Setiawan; Cita S Amalia; Dwi AA Nugrahaningsih; Dwi Indaryati; Dyah A Puspitarani; Edwin W. Daniwijaya; Eggi Arguni; Endah Supriyati; Gunadi; Hana F Hanifin; Hendra Wibawa; I Putu Aditio A; Indarto Sulistiyono; Irene Tania; Kristy Iskandar; Ludhang P. Rizki; Marcellus; Mohamad S. Hakim; Nungki Anggorowati; Pramesti G Dewi; Siswanto; Susan Simanjaya; Titik Nuryastuti; Tri Wibawa

Afiahayati; Alvina A Setiawan; Dwi AA Nugrahaningsih; Dwi Indaryati; Edwin W. Daniwijaya; Eggi Arguni; Endah Supriyati; Gunadi; Hendra Wibawa; Indarto Sulistiyono; Khanza a Vujira; Kristy Iskandar; Ludhang P. Rizki; Marcellus; Mohamad S. Hakim; Nungki Anggorowati; Pramesti G Dewi; Siswanto; Titik Nuryastuti; Tri Wibawa

EPI_ISL_2954064	RSUD Panembahan Senopati, Bantul	Genetics Working Group (Pokja Genetik) Faculty of Medicine, Public Health and Nursing Universitas Gadjah Mada (FK-KMK UGM); Disease

Investigation Center Wates Ministry of Agriculture Indonesia; Department of Microbiology FK-KMK UGM; Laboratorium Diagnostik Yayasan Tahija World Mosquito Program (WMP) Yogyakarta Center for Tropical Medicine FK-KMK UGM; Integrated Research Center FK-KMK UGM; Department of Computer

Afiahayati; Dwi AA Nugrahaningsih; Dwi Indaryati; Edwin W. Daniwijaya; Eggi Arguni; Endah Supriyati; Gunadi; Hana F Hanifin; Hendra Wibawa; Indarto Sulistiyono; Irene Tania; Kristy Iskandar; Ludhang P. Rizki; Marcellus; Mohamad S. Hakim; Nungki Anggorowati; Pramesti G Dewi; Siswanto; Titik Nuryastuti; Tri Wibawa

EPI_ISL_2502616, EPI_ISL_2502627, EPI_ISL_2502628, EPI_ISL_2502629, EPI_ISL_2502667, EPI_ISL_2502687, EPI_ISL_2502717, EPI_ISL_2502763, EPI_ISL_2502764, EPI_ISL_2502765, EPI_ISL_2506039, EPI_ISL_2506040, EPI_ISL_2506041, EPI_ISL_2534078, EPI_ISL_2534079, EPI_ISL_2534082, EPI_ISL_2534289, EPI_ISL_2534338, EPI_ISL_2534346, EPI_ISL_2534376, EPI_ISL_2534377, EPI_ISL_2534379, EPI_ISL_2534454, EPI_ISL_2534456, EPI_ISL_2534457, EPI_ISL_2534458, EPI_ISL_2534490, EPI_ISL_2534518

see above	RSUD dr. LOEKMONO HADI	Genetics Working Group (Pokja Genetik) Faculty of Medicine, Public Health and Nursing Universitas Gadjah Mada (FK-KMK UGM); Disease

Investigation Center Wates Ministry of Agriculture Indonesia; Department of Microbiology FK-KMK UGM; Laboratorium Diagnostik Yayasan Tahija World Mosquito Program (WMP) Yogyakarta Center for Tropical Medicine FK-KMK UGM; Integrated Research Center FK-KMK UGM; Department of Computer Science and Electronics FMIPA UGM; RSUP Dr. Sardjito

EPI_ISL_2955607	RSUP Dr Sardjito	Genetics Working Group (Pokja Genetik) Faculty of Medicine, Public Health and Nursing Universitas Gadjah Mada (FK-KMK UGM); Disease

Investigation Center Wates Ministry of Agriculture Indonesia; Department of Microbiology FK-KMK UGM; Laboratorium Diagnostik Yayasan Tahija World Mosquito Program (WMP) Yogyakarta Center for Tropical Medicine FK-KMK UGM; Integrated Research Center FK-KMK UGM; Department of Computer Science and Electronics FMIPA UGM; RSUP Dr. Sardjito

EPI_ISL_2943173	RSUP Dr. Sardjito	Genetics Working Group (Pokja Genetik) Faculty of Medicine, Public Health and Nursing Universitas Gadjah Mada (FK-KMK UGM); Disease

Investigation Center Wates Ministry of Agriculture Indonesia; Department of Microbiology FK-KMK UGM; Laboratorium Diagnostik Yayasan Tahija World Mosquito Program

EPI_ISL_2955606	RSUP Sardjito	Genetics Working Group (Pokja Genetik) Faculty of Medicine, Public Health and Nursing Universitas Gadjah Mada (FK-KMK UGM); Disease

Investigation Center Wates Ministry of Agriculture Indonesia; Department of Microbiology FK-KMK UGM; Laboratorium Diagnostik Yayasan Tahija World Mosquito Program (WMP) Yogyakarta Center for Tropical Medicine FK-KMK UGM; Integrated Research Center FK-KMK UGM; Department of Computer Science and Electronics FMIPA UGM; RSUP Dr. Sardjito

Afiahayati; Alvina A Setiawan; Cita S Amalia; Dwi AA Nugrahaningsih; Dwi Indaryati; Dyah A Puspitarani; Edwin W. Daniwijaya; Eggi Arguni; Endah Supriyati; Gunadi; Hana F Hanifin; Hendra Wibawa; Indarto Sulistiyono; Irene Tania; Khanza a Vujira; Kristy Iskandar; Ludhang P. Rizki; Marcellus; Mohamad S. Hakim; Nungki Anggorowati; Pramesti G Dewi; Siswanto; Susan Simanjaya; Titik Nuryastuti; Tri Wibawa

. Marcellus; Afiahayati; Bambang Sigit Riyanto; Dwi AA Nugrahaningsih; Edwin W. Daniwijaya; Eggi Arguni; Eko Budiono; Endah Supriyati; Gunadi; Hana F Hanifin; Hendra Wibawa; Heni Retnowulan; Ika Trisnawati; Ira Puspitawati; Irene Tania; Kristy Iskandar; Ludhang P. Rizki; Mohamad S. Hakim; Munawar Gani; Nungki Anggorowati; Nur Imma Fatimah Harahap; Nur Rahmi Ananda; Osman Sianipar; Riat El Khair; Satria Maulana; Siswanto; Sumardi; Titik Nuryastuti; Tri Wibawa; Umi Solekhah Intansari; Yunika Puspadewi; Elizabeth Henny Herningtiyas

. Marcellus; Afiahayati; Bambang Sigit Riyanto; Dwi AA Nugrahaningsih; Edwin W. Daniwijaya; Eggi Arguni; Eko Budiono; Endah Supriyati; Gunadi; Hana F Hanifin; Hendra Wibawa; Heni Retnowulan; Ika Trisnawati; Ira Puspitawati; Irene Tania; Kristy Iskandar; Ludhang P. Rizki; Mohamad S. Hakim; Munawar Gani; Nungki Anggorowati; Nur Imma Fatimah Harahap; Nur Rahmi Ananda; Osman Sianipar; Riat El Khair; Satria Maulana; Siswanto; Sumardi; Titik Nuryastuti; Tri Wibawa; Umi Solekhah Intansari; Yunika Puspadewi; Elizabeth Henny Herningtiyas

. Marcellus; Afiahayati; Alvina A Setiawan; Bambang Sigit Riyanto; Dwi AA Nugrahaningsih; Edwin W. Daniwijaya; Eggi Arguni; Eko Budiono; Endah Supriyati; Gunadi; Hendra Wibawa; Heni Retnowulan; Ika Trisnawati; Ira Puspitawati; Khanza a Vujira; Kristy Iskandar; Ludhang P. Rizki; Mohamad S. Hakim; Munawar Gani; Nungki Anggorowati; Nur Imma Fatimah Harahap; Nur Rahmi Ananda; Osman Sianipar; Riat El Khair; Satria Maulana; Siswanto; Sumardi; Titik Nuryastuti; Tri Wibawa; Umi Solekhah Intansari; Yunika Puspadewi; Elizabeth Henny Herningtiyas
